# Androgen receptor pathway activity assay for sepsis diagnosis and prediction of favorable prognosis

**DOI:** 10.1101/2021.07.07.21259768

**Authors:** Wilbert Bouwman, Wim Verhaegh, Anja van de Stolpe

## Abstract

**Introduction:** Sepsis is a life-threatening complication of a bacterial infection. It is hard to predict which patients with a bacterial infection will develop sepsis, and accurate and timely diagnosis, as well as assessment of prognosis, is difficult. Aside from antibiotics-based treatment of the causative infection and circulation-supportive measures, treatment options remain limited. Better understanding of the immuno-pathophysiology of sepsis may lead to improved diagnostic and therapeutic solutions.

Functional activity of the innate (inflammatory) and adaptive immune response is controlled by a dedicated set of cellular signal transduction pathways in the various immune cell types. To develop an immune response-based diagnostic assay for sepsis and provide novel therapeutic targets, signal transduction pathway activities were analyzed in whole blood samples from patients with sepsis.

**Methods:** A validated and previously published set of signal transduction pathway (STP) assays, determining immune cell function, was used to analyze public Affymetrix expression microarray data from clinical studies containing data from pediatric and adult patients with sepsis. STP assays enable quantitative measurement of STP activity on individual patient sample data, and were used to calculate activity of androgen receptor (AR), estrogen receptor (ER), JAK-STAT1/2, JAK-STAT3, Notch, Hedgehog, TGFβ, FOXO-PI3K, MAPK-AP1, and NFκB signal transduction pathways.

**Results:** Activity of AR and TGFβ pathways was increased in children and adults with sepsis. Using the mean plus two standard deviations of normal pathway activity (in healthy individuals) as threshold, diagnostic assay parameters were determined. For diagnosis of pediatric sepsis, the AR pathway assay showed high sensitivity (77%) and specificity (97%), with a PPV of 99% and NPV of 50%. For prediction of favorable prognosis (survival), PPV was 95%, NPV was 21%. The TGFβ pathway activity assay performed slightly less for diagnosing sepsis, with a sensitivity of 64% and specificity of 98% (PPV 99%, NPV 39%).

**Conclusion:** The AR and TGFβ pathways have an immunosuppressive role, suggesting a causal relation between increased pathway activity and sepsis immunopathology. STP assays have been converted to qPCR assays for further evaluation of clinical utility for sepsis diagnosis and prediction of prognosis, as well as for prediction of risk at developing sepsis in patients with a bacterial infection. STPs may present novel therapeutic targets in sepsis.

## Introduction

Sepsis is a life-threatening infection in which the immune response is dysregulated resulting in multi-organ dysfunction or failure [1]. Sepsis is generally a complication of severe bacterial infection and characterized by a systemic inflammatory response leading to septic shock. Mortality rates range between 25% to 30% for severe sepsis and 40% to 70% for septic shock [2],[3].

Aside from antibiotics and supportive measures to maintain blood circulation of internal organs, no treatments have proven to be effective, although it cannot be excluded that some treatments may benefit a subset of patients who so far cannot be identified [1]. One reason for failure to develop effective treatments is the heterogeneity among sepsis patients, that is, variation in underlying medical conditions and use of drugs, as well as genetic variations influencing the immune response in an individual patient.

Detailed assessment of the functional immune response may enable a personalized treatment approach and improve treatment efficacy. Diagnostic assessment of immune function is currently limited to routine blood measurements, such as numbers of immune cells and inflammation markers like C-reactive protein (CRP), but is not informative on the functional state of the various types of immune cells, which is responsible for the abnormal immune response in a sepsis patient.

The functional state of immune cells is determined by a small number of so-called cellular signal transduction pathways (STPs) [4],[5],[6],[7],[8]. Recently, novel assays have been developed to quantitatively measure activity of STPs in cell and tissue samples, including blood samples [9],[10],[11],[12]. Measuring combined activity of these STPs is expected to enable quantitative assessment of the innate and adaptive immune response in an individual patient [7],[13]. In this study, STP analysis was performed on publicly available gene expression data from multiple clinical sepsis studies. Measurement of activity of the androgen receptor (AR) pathway, and to a lesser extent the TGFβ pathway, in a whole blood sample is shown to have value for sepsis diagnosis and prediction of prognosis in a sepsis patient, and may lead to novel personalized treatment options. Measurement of STP activity to identify patients with a bacterial infection who are at high risk to develop sepsis is discussed.

## Methods

### STP assays to determine activity of androgen receptor (AR), estrogen receptor (ER), FOXO-PI3K, JAK-STAT1/2, JAK-STAT3, Notch, Hedgehog (HH), TGFβ, and NFκB pathways in blood cells

Development and validation of assays to quantify STP activity have been described before [9]– [11],[12]. In brief, target genes of transcription factors of the respective signal transduction pathways were identified, and a Bayesian network computational model was created for interpretation of measured mRNA levels of the pathway target genes to generate a quantitative pathway activity score (PAS). PAS are presented on a log2 odds scale as described [14],[15].

### Affymetrix expression microarray data analysis

For analysis of clinical studies, STP assays were performed on public Affymetrix HG-U133 Plus2.0 microarray expression datasets from previously published clinical studies (deposited in the GEO database [16]). Quality control (QC) was performed on Affymetrix data of each individual sample prior to STP analysis, as described before [10], using available R packages [17],[18]. Samples that failed QC were removed prior to data analysis.

A summary of clinical datasets used in this study is shown in Supplementary Table 1. All studies provided Affymetrix data from whole blood samples. Duplicate sample data between datasets of different studies were removed when such datasets were combined into one analysis or calculation, such as for determining normal pathway activity thresholds and for sensitivity and specificity calculations. For these purposes, 8 duplicates from GSE26640, 28 duplicates from GSE8121, 10 duplicates from GSE9692, 48 duplicates from GSE13904, and 1 duplicate from GSE26378 were removed (specified in Supplementary Table 1).

**Table 1.** Performance parameters of the AR pathway assay for pediatric sepsis diagnosis and prediction of survival.

|  | **Sepsis diagnosis** | **Sepsis survival** | **Sepsis non survival** |
| --- | --- | --- | --- |
|  | **AR PAS above**<br>**normal range**<br>**n=492** | **AR PAS in normal**<br>**range**<br>**n=255** | **AR PAS above normal**<br>**range**<br>**n=255** |
| **Sepsis** |  |  |  |
| **Sensitivity** | 77% | 25% | 93% |
| **Specificity** | 97% | 93% | 25% |
| **PPV** | 99% | 95% | 21% |
| **NPV** | 50% | 21% | 95% |
| **Fisher's Exact Test** | p-value < 2.2e-16 | p-value = 0.005023 | p-value = 0.005023 |

### Interpretation of signal transduction pathway activity scores (PAS)

An important and unique advantage of the STP assays is that in principle they can be performed on all cell types. A few considerations to bear in mind when interpretating log2 odds PAS, as described before [19], are as follows:

1. On the same sample, log2 odds PAS cannot be compared between different signaling pathways, since each of the signaling pathways has its own range in log2 odds activity scores [10].
2. The log2 odds range for pathway activity (minimum to maximum activity) may vary depending on cell type. Once the range has been defined using samples with known pathway activity, on each new sample the absolute value can be directly interpreted against that reference. If the range has not been defined, only differences in log2 odds activity score between samples can be interpreted.
3. PAS are highly quantitative, and even small differences in log2 odds PAS can be reproducible and meaningful.
4. A negative log2 odds ratio does not necessarily mean that the pathway is inactive.

### Statistics

Boxplots were made using the Python data visualization library function *seaborn*. Statistical annotations were created using the Python package statannot [20], [21]. Two sided Mann-Whitney-Wilcoxon testing was used to compare PAS across groups. For paired testing (dataset GSE95233) a one-sided t-test was used. p-values are indicated in the figures. In accordance with current consensus regarding the use of statistical parameters, a p-value of 0.01 (indicated as double asterisk in the figures) was considered significant [22]. Receiver Operating Characteristics (ROC) curves and Area Under Curve (AUC) was calculated in R. Thresholds for diagnosis and prognosis classification were based on the normal range for pathway activity in healthy individuals, as determined by mean +/- two standard deviations (SD) of pathway activity of healthy individuals. Based on these thresholds, sensitivity, specificity, PPV and NPV were determined, and Fisher exact tests were used to compare proportions between groups.

## Results

### Measuring STP activity in whole blood samples from pediatric and adult patients with sepsis

STP activity scores from whole blood samples for the AR, ER, MAPK-AP1, FOXO-PI3K, JAK-STAT1/2, JAK-STAT3, Notch, Hedgehog, TGFβ, and NFκB pathways were measured using target gene expression data from clinical studies on children (Supplementary Figures S1-S6) and adults with sepsis and septic shock (Supplementary Figures S7-S8). An overview of all included datasets is available in Supplementary Table 1.

### AR and TGFβ signal transduction pathway activity scores are higher in patients with sepsis/septic shock

Activity of the AR pathway, and with exception of clinical study GSE57065 also of the TGFβ pathway, were consistently and significantly increased in pediatric and adult patients with sepsis and septic shock (Figure 1-4). No consistent differences in AR and TGFβ pathway activity scores were observed between men and women (Figure 1F, 2F, Supplementary Figure S6), between patients categorized as “sepsis” versus “septic shock” (Figure 1D, 2D, Supplementary Figure S4), nor between patients categorized as “SAPSII-low” and “SAPSII-high” (Figure 3A, 4A, Supplementary Figure S7). No difference was found between patients with gram-negative and gram-positive bacterial infections (clinical study datasets GSE4607 and GSE9692; data not shown).

**Figure 1:**
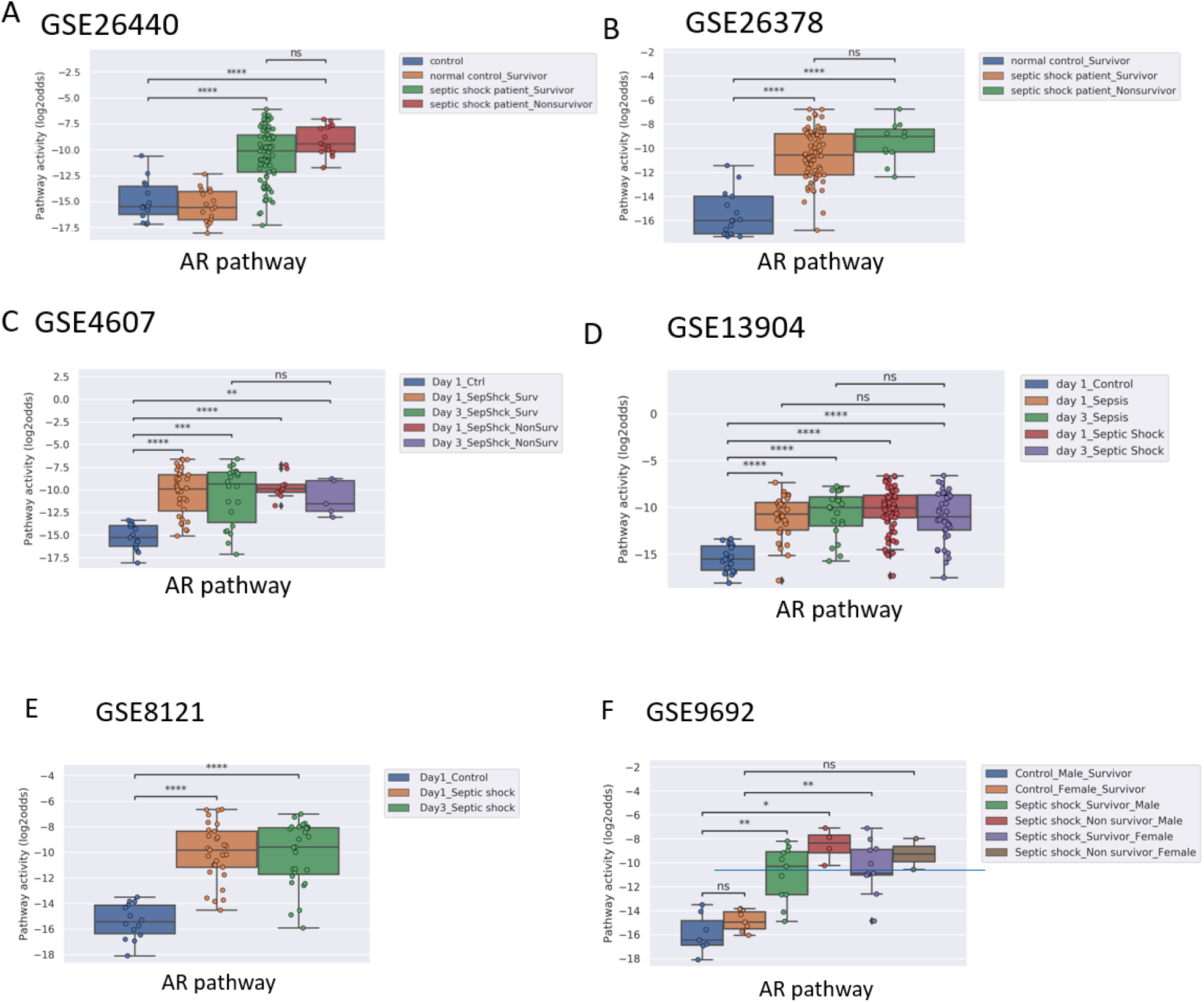
AR pathway activity scores (PAS) of Affymetrix datasets of whole blood samples from children (<10 years of age). Samples were obtained within 24 hours of initial presentation to the pediatric intensive care unit (PICU) with sepsis (all datasets). Indicated are when applicable: survivors and non-survivors (A,B,C,F); time (Day) on PICU (C); and gender (F).Control patients were recruited from the participating institutions using the following exclusion criteria: a recent febrile illness (within 2 weeks), recent use of anti-inflammatory medications (within 2 weeks), or any history of chronic or acute disease associated with inflammation. **A)** Dataset GSE26440 [60]. **B)** GSE26378 [60]. **C)** Dataset GSE4607 [61]. **D)** Dataset GSE13904 [27]. **E)** Dataset GSE8121 [28] **F)** Dataset GSE9692 [62]. AR PAS on Y-axis on a log2 odds scale. Two sided Mann–Whitney–Wilcoxon statistical tests were performed; p-values are indicated in the figures as **p < 0.01, ***p < 0.001, ****p < 0.0001 or ns (not significant).

**Figure 2:**
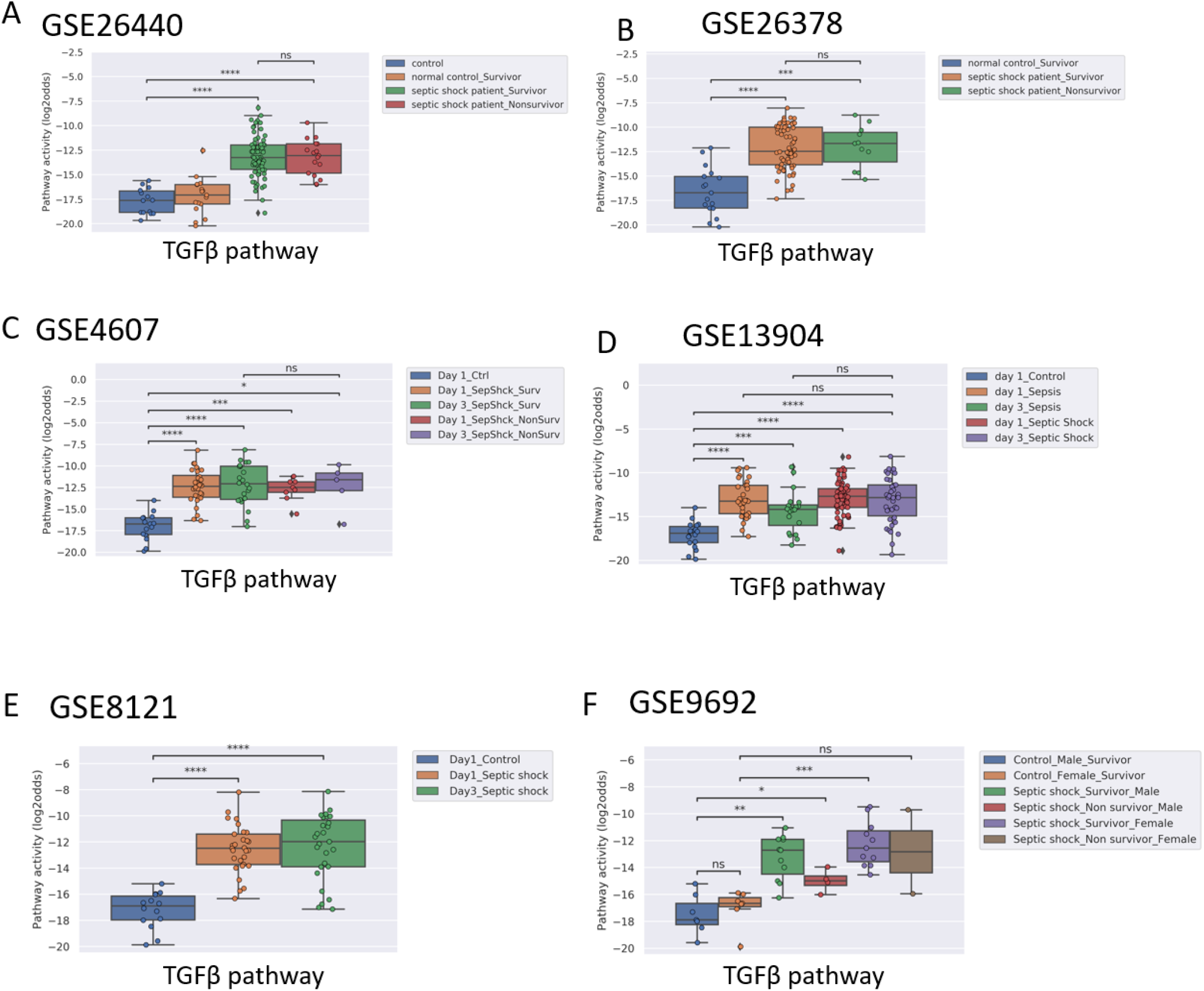
TGFβ PAS, same datasets as in figure 1.

**Figure 3:**
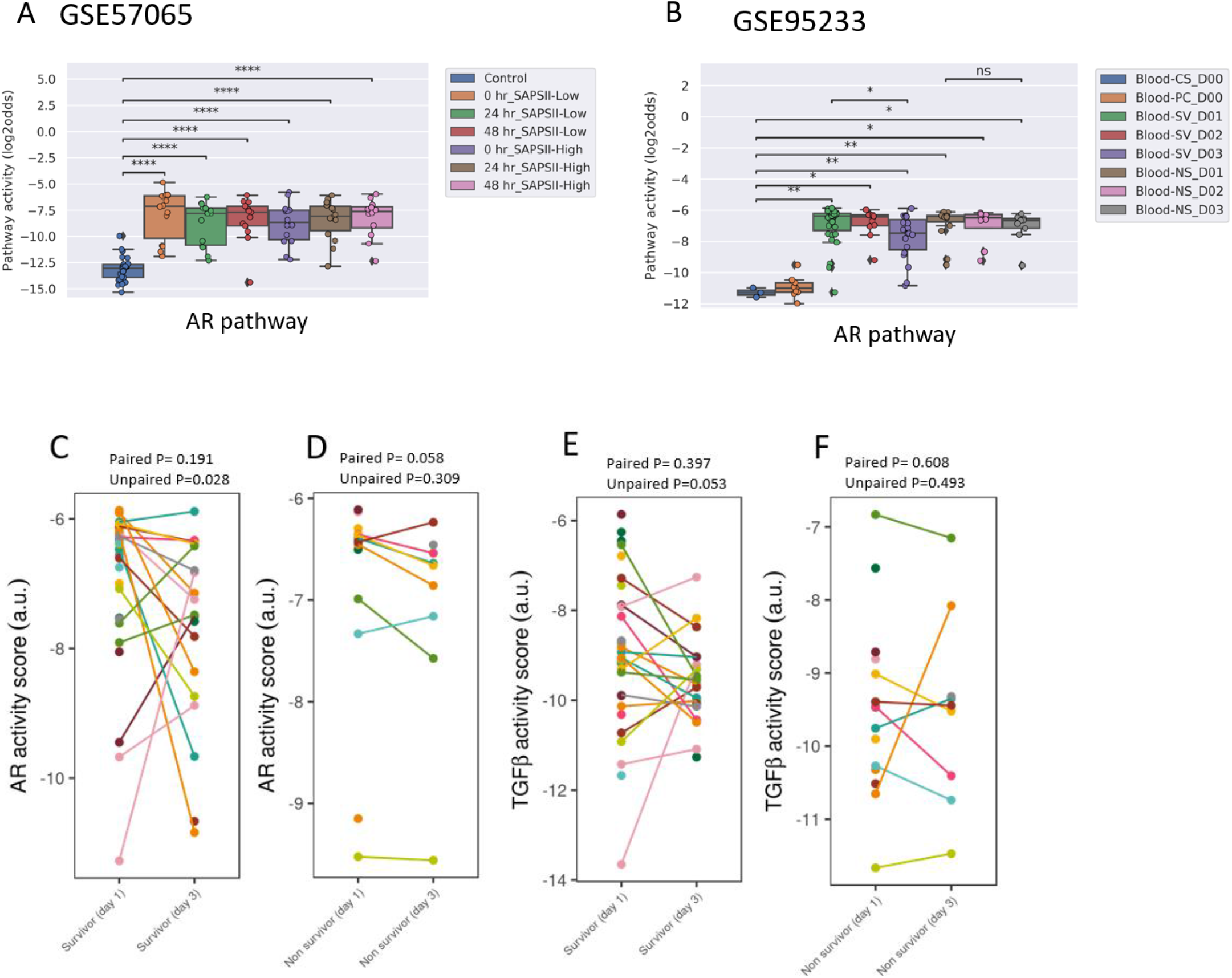
AR PAS of datasets of whole blood samples from adults with sepsis and healthy individuals. **A)** Dataset GSE57065 [23], containing ICU patients enrolled at the onset of septic shock. Blood samples collected within 30 minutes, 24 and 48 hours after diagnosis of sepsis. Sepsis patients were divided into SAPSII-low and SAPSII-high groups according to the median of SAPSII score (SAPSII scores of <45 and >45, respectively). **B)** Dataset GSE95233 [24] containing ICU patients enrolled at diagnosis of septic shock. Blood samples collected at admission, and at day 2 (D2) or day 3 (D3). SV indicates survivor, NS indicates non-survivor sepsis patient. CS indicates control healthy individual; PC indicates patient control. **C-E)** Dataset GSE95233 [24]. AR (C,D) and TGFβ (E,F) PAS for sepsis survivor versus non-survivor patients, at day 1 and day 3 after diagnosis. Patient sample are connected by lines. AR PAS on Y-axis on log2 odds scale. For A,B two sided Mann–Whitney–Wilcoxon statistical tests; for C,D a one sided paired and unpaired t-test. p-values are indicated as ****p < 0.0001 or ns (not significant).

Two adult sepsis clinical studies (GSE57065 [23]; (GSE95233 [24]) allowed investigation of STP activity at several time points after disease onset. Blood samples were collected within 30 minutes, and at 24 and 48 hours after septic shock onset (Figure 3A, 4A, Supplementary Figures S7) or were collected at Day 1, Day 2, or Day 3 (Figure 3B, Supplementary Figures S8, S11). AR pathway activity scores were already significantly increased at first measurement and remained increased over the next days.

### Normal range AR pathway activity in patients with sepsis may be indicative for survival

In pediatric sepsis patients who survived, AR (but not TGFβ) pathway activity showed a trend towards lower PAS, within the PAS range of healthy controls (Figures 1A, 1B, 1C and 1F; 2A, 2B, 2C). Only one adult sepsis study allowed investigation of the relation between survival and STP activity (GSE95233 [24], Figure 3B-F, Supplementary Figures S8, S11). AR pathway activity tended to be lower for sepsis survivors at day three after diagnosis, and tended to decrease between Day 1 and Day 3 of sepsis in survivor patients and (Figures 3B-D, S11). For the TGFβ pathway this was not found (Figures 4B and 3E-F, S11). Although not significant, results from these independent studies suggest that a normal range AR pathway activity may be favorable in patients with sepsis.

**Figure 4:**
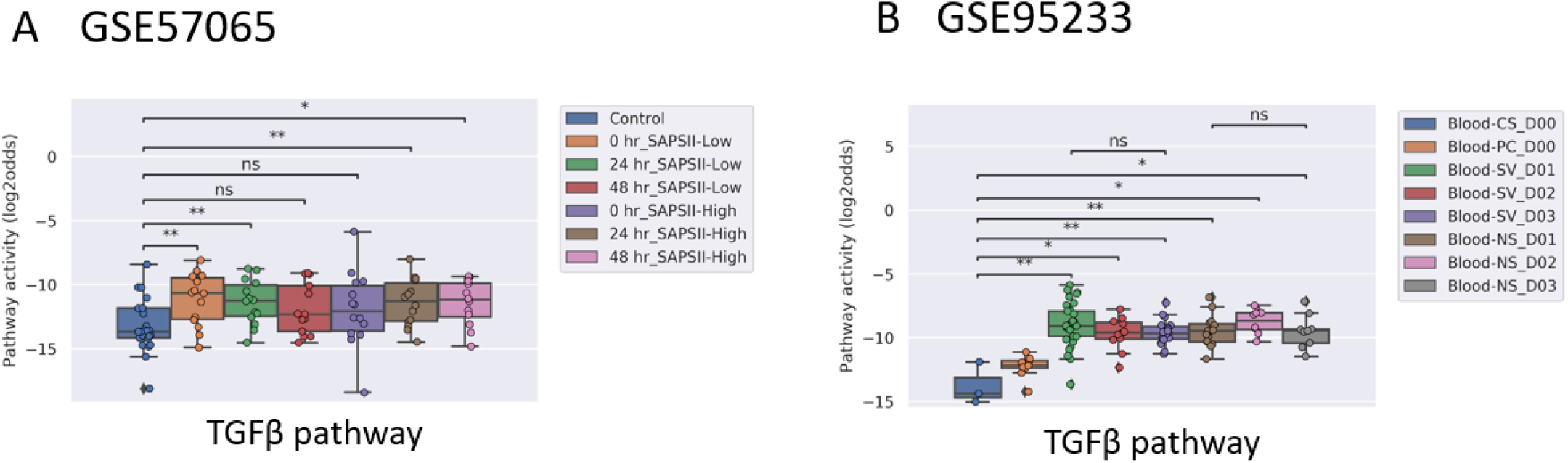
TGFβ PAS, same datasets as in Figure 3.

### Activity of other signaling pathways (Figures S1-S8)

ER pathway PAS were low and increased slightly in patients with sepsis, but the increase was not significant in dataset GSE95233 (pediatrics) and GSE9692 (adults). MAPK-AP1, FOXO-PI3K, JAK-STAT1/2, JAK-STAT3, NFκB and Notch signaling pathway PAS were either not significantly increased in sepsis patients, or not consistently increased across multiple clinical studies. Activity of the Hedgehog pathway tended to be lower in sepsis patients, only meeting statistical requirements in the adult sepsis studies (GSE57065 and GSE95233) (Supplementary Figures S7 and S8).

### Defining an upper threshold for normal AR and TGFβ pathway activity in whole blood samples

To enable determination of STP assay performance parameters for diagnostic use in sepsis patients, normal STP PAS ranges in whole blood samples were determined. Eight datasets contained samples from healthy individuals (GSE26440 [23], GSE26378 [25], GSE4607 [26], GSE13904 [27], GSE8121[28] and GSE9692 [29]) (Supplementary Table S1).

Mean AR PAS in healthy individuals was -15.37 (log2 odds scale, SD 1.52) for children (n=93) and -12.44 (log2 odds scale, SD 1.44) for adults (n=37) (Supplementary Table S2). Combining healthy pediatric and adult sample data (n=130), mean AR PAS was -14.5 (log2 odds scale, SD 2.0). No significant differences between men and women were found. Using this information, normal (healthy) STP PAS thresholds were determined for each STP, based on the mean STP PAS score in healthy controls +/- 2 standard deviations (SD) for combined datasets (Supplementary Tables S2) and for individual datasets (Supplementary Table S4). The upper AR PAS threshold for normal was calculated as the mean + 2SD, resulting in a threshold for pediatric patients of - 12.33 (log2 odds scale), for adults -9.57 (log2 odds scale), and for pediatric and adults combined -10.54 (log2 odds scale). For the TGFβ pathway mean PAS in healthy individuals was -17.05 (log2 odds scale, SD 1.63) for children and -13.01 (log2 odds scale, SD 1.83) for adults (Supplementary Table S2). Combining healthy pediatric and adult sample data, mean TGFβ PAS was -15.9 (log2 odds scale, SD 2.49).

Applying the pediatric upper threshold for normal AR PAS on the pediatric study data showed that only 3 out of 45 (7%) children in the non-survivor groups had an AR pathway activity in the normal range, while 53 out of 210 (25%) survivor children had AR pathway activity in the normal range (Fisher exact test p = 0.005). For the TGFβ pathway the percentages were respectively 31 and 32. Thus, AR PAS in the normal range was associated with sepsis survival, at least in children (Figures 1A-C, F, Figures 2A-C, F). We proceeded with calculating AR and TGFβ pathway assay performance parameters.

### Sensitivity and specificity of AR and TGFβ pathway activity assays for pediatric sepsis diagnosis and prognosis prediction

Using the pediatric upper threshold of the normal range of AR and TGFβ PAS, sensitivity, specificity, PPV, and NPV for diagnosis and prediction of survival in a pediatric sepsis patient were calculated (Table 1, Table 2, Supplementary Tables 5 and 6), and a ROC curve was generated (Figure 5). For the same parameters for the other STPs, see Supplementary Figures S9 and S10. For sepsis diagnosis, AR pathway assay showed both a high sensitivity (77%) and specificity (97%), with a PPV of 99% and a NPV of 50%. AUC in the ROC curve was 0.94 for sepsis diagnosis. For prediction of favorable (survivor) prognosis, the PPV was 95%, indicating that the assay was highly specific (93%) in identifying survivor patients. The NPV was 21%, indicative of low sensitivity (25%) in predicting survivor patients. The TGFβ assay performed less, with a sensitivity for diagnosing sepsis of respectively 64% and specificity of 98%; all other STP assays performed less well (Supplementary Figure S9).

**Table 2.**
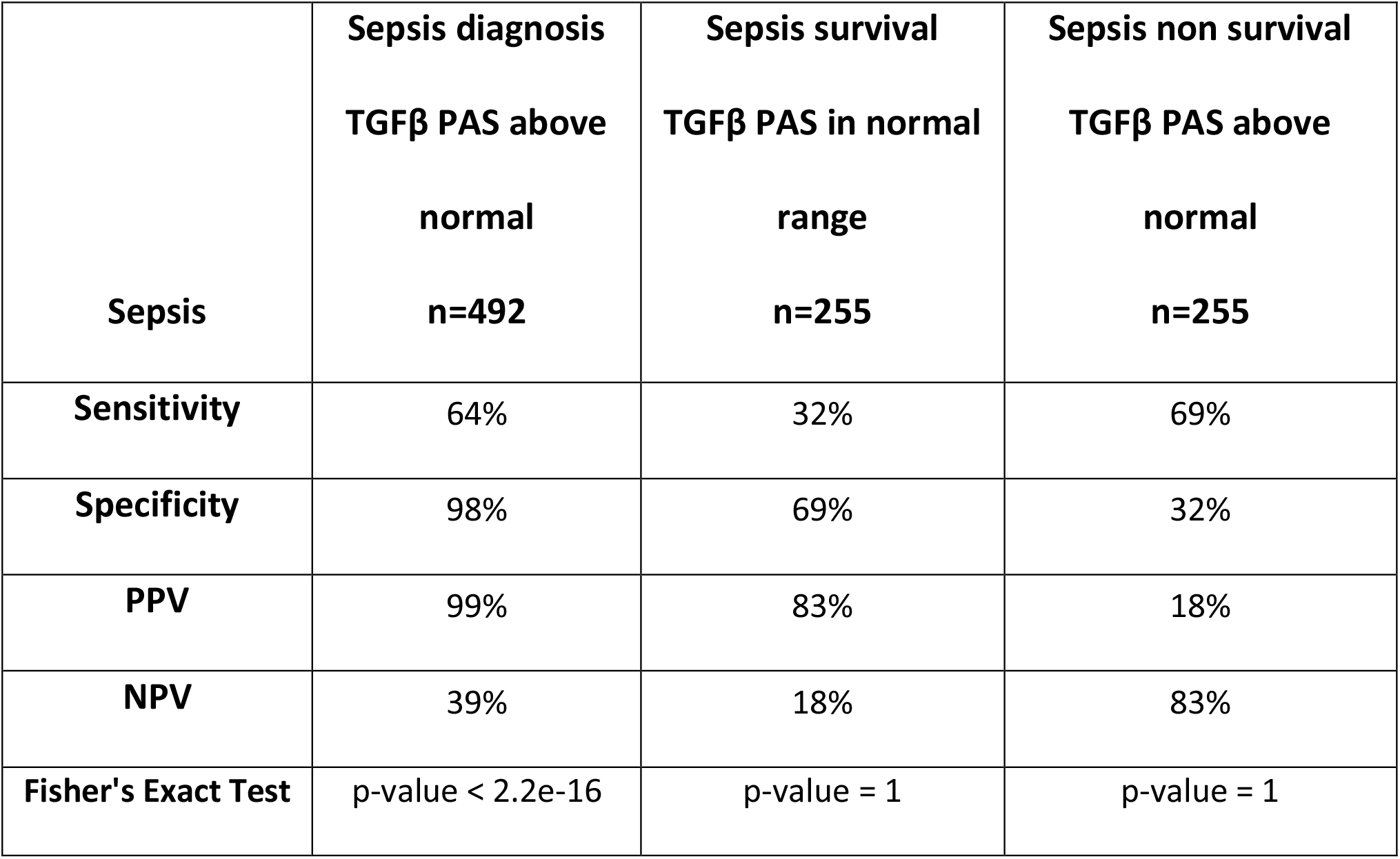
Performance parameters of the TGFβ pathway assay for pediatric sepsis diagnosis and prediction of survival.

|  | **Sepsis diagnosis** | **Sepsis survival** | **Sepsis non survival** |
| --- | --- | --- | --- |
|  | **TGFβ PAS above**<br>**normal**<br>**n=492** | **TGFβ PAS in normal**<br>**range**<br>**n=255** | **TGFβ PAS above**<br>**normal**<br>**n=255** |
| **Sepsis** |  |  |  |
| **Sensitivity** | 64% | 32% | 69% |
| **Specificity** | 98% | 69% | 32% |
| **PPV** | 99% | 83% | 18% |
| **NPV** | 39% | 18% | 83% |
| **Fisher's Exact Test** | p-value < 2.2e-16 | p-value = 1 | p-value = 1 |

**Figure 5.**
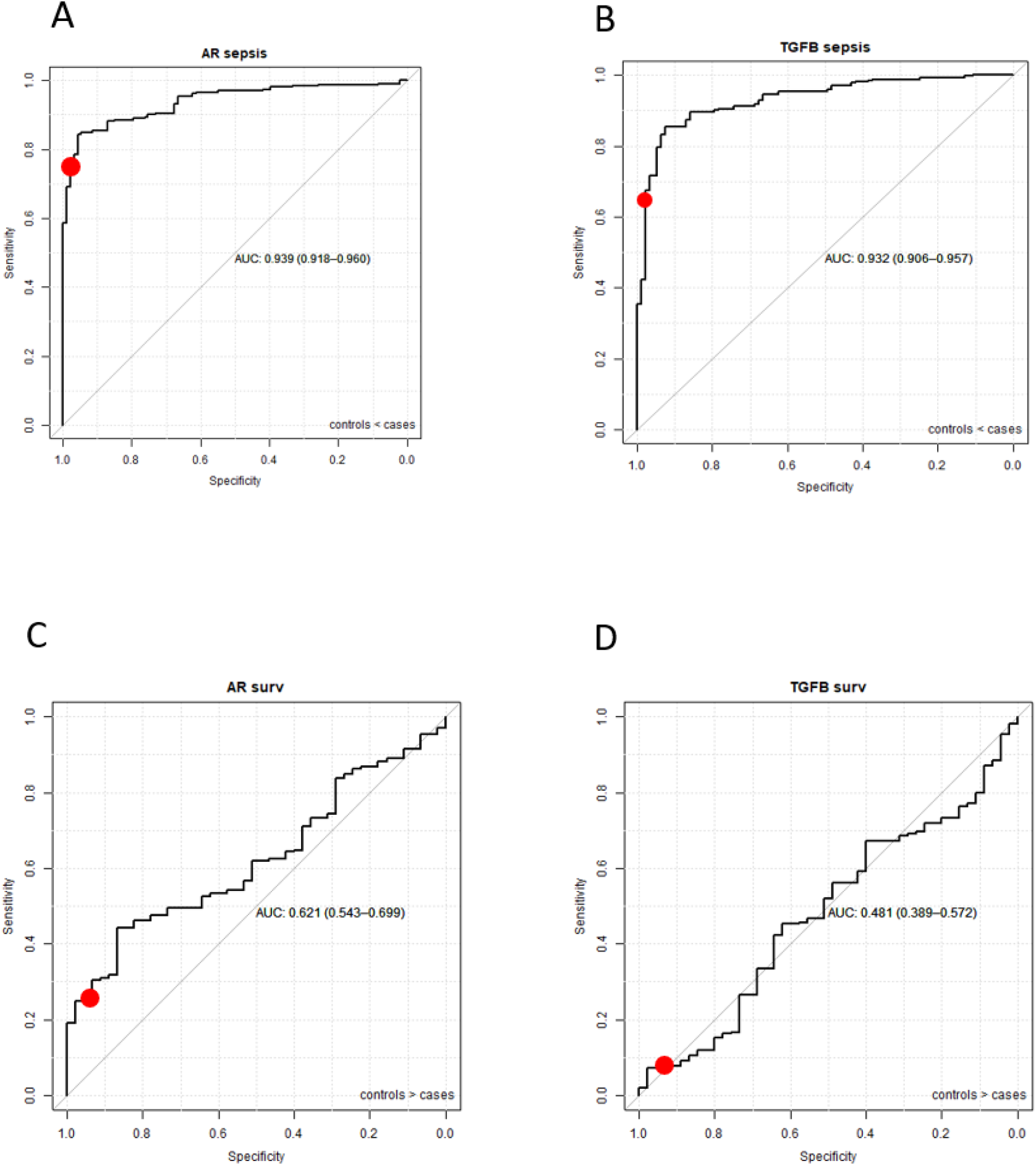
ROC curves (and AUC) of AR and TGFβ pathway activity assays for sepsis diagnosis (A,B) and survival prediction (C,D). Healthy controls and sepsis patients (sepsis and sepsis shock combined) patients from datasets GSE26440 [60], GSE26378 [60], GSE4607 [61], GSE13904 [27], GSE8121[30] and GSE9692 [62] are included (duplicate samples excluded). The red dot represents sensitivity and specificity when the upper threshold is the mean plus 2 SD of control (healthy individuals) samples.

### Comparison of Affymetrix data analysis results described in the original clinical study analysis publications with results of here described STP analysis

For each Affymetrix dataset for which we performed STP analysis, bioinformatics tools used for data analysis as reported in the associated publication have been listed, together with reported functional gene annotations and/or identified “pathways” as defined by the used tools (Ingenuity, PANTHER, D.A.V.I.D., ToppGene) (see Supplementary information). In all analyses, groups of patient samples had been compared with respect to differential gene expression patterns. In contrast to the current study, individual samples had not been analyzed. An association between sepsis/septic shock and signal transduction pathways had been identified using Ingenuity Pathways Analysis and PANTHER. The NFκB pathway (termed NFκB or TLR) [25],[30],[29],[26], the MAPK pathway (termed “p380MAPK, PDGF”) [30],[29],[31],[26], the JAK-STAT3 pathway (termed “IL6, IL10”) [30],[29],[31],[26], and the PI3K-FOXO pathway (termed “Integrin signaling/Insulin signaling/IGF1 signaling”) [30], were identified as associated with sepsis, but without information on activity state of the pathways. Hormonal AR and ER pathways were not mentioned and neither was the TGFβ pathway.

## Discussion

Using a previously reported Signal Transduction Pathway (STP) assay platform, we quantified activity of the most important signal transduction pathways that determine immune cell function, using RNA expression data from whole blood samples of previously published clinical pediatric and adult sepsis studies [9],[12],[10],[8],[11],[15].

### Several signaling pathway activities are increased in whole blood from patients with sepsis: potential for diagnostic use

Activity of the AR pathway, and to a lesser extent of the TGFβ signaling pathway, was increased in sepsis, while a trend towards higher activity was observed for the MAPK-AP1, ER, NFκB and JAK-STAT3, and towards lower activity of the Hedgehog pathway, while activity of the JAK-STAT1/2 pathway did not change. Only the AR pathway assay could to some extent identify survivor sepsis patients. This assay showed high sensitivity and specificity for sepsis diagnosis, and high specificity for prediction of favorable (survival) prognosis in children with sepsis.

In sepsis, the immune system plays a crucial role, with both pro-inflammatory and anti-inflammatory mechanisms. A subset of patients with sepsis rapidly displays signs of immunosuppression and inflammation associated with worse prognosis [32],[33]. In line with our observations, for AR, ER, TGFβ, NFκB, and JAK-STAT3 pathways various functional roles in the immune response have been described before, with a distinct immunosuppressive role for AR and TGFβ pathways and an inflammatory role for NFκB and JAK-STAT3 pathways [34], [35],[36],[37], [32],[33], [38], [39], [40], [41].

The lack of change in JAK-STAT1/2 pathway activity which we observed is in agreement with our earlier findings that PAS scores only increase in virally, but not bacterially, infected patients, at least when measured in Peripheral Blood Mononuclear Cell (PBMC) samples [8]. For the AR, ER, TGFβ, and NFκB pathways putative roles in sepsis have been described, generally in relation to specific immune cell types [42],[43],[44],[45],[46]. Immune cell-specific regulation of pathway activity is a likely explanation for lack of sufficient diagnostic power of som of the measured STPs when measured in whole blood samples with its intrinsic heterogeneity in immune cell composition.

### Is there a causal relation between AR pathway activity and sepsis?

Despite the disadvantage of measuring in heterogeneic whole blood samples, the diagnostic performance of AR and TGFβ assays for sepsis was good. With respect to the TGFβ pathway, either decreased or elevated levels of various TGFβ pathway ligands have been reported in sepsis patients, while uncertainty remains with respect to the causal role of this pathway [46]. Multiple findings support a causal role for the AR pathway in sepsis. The AR signaling pathway is in principle activated by the presence of testosterone ligand, binding to a pocket in the AR transcription factor protein to induce RNA transcription of target genes [6]. The AR protein is expressed in a wide variety of innate and adaptive immune cells including neutrophils, macrophages, mast cells, monocytes, megakaryocytes, B cells, and T cells, indicating that the AR signaling pathway can be activated in these blood cell types [44]. Monocytes are one of the key cell types that play a pathogenic role in sepsis, and make up around 10% of whole blood cells [47],[48]. Interestingly and in line, men have shown to be more susceptible to developing sepsis than women [49]. Some studies suggest that the AR pathway may be a useful drug target in sepsis: testosterone blockade in patients with hemorrhage improved prognosis if subsequently sepsis developed [50] and AR pathway blockade reduced mortality in a preclinical mouse model for sepsis [42],[51]. While suggestive of a role for the AR pathway in the pathophysiology of sepsis, we are not aware of any clinical studies investigating AR pathway targeted drugs in human sepsis patients.

### AR pathway assay as a diagnostic assay for sepsis

Current diagnosis of sepsis is based on assessment of multiple clinical symptoms and biomarkers, but timely diagnosis and even more so, prediction of prognosis of a sepsis patient, remain a clinical challenge. Despite the limitation of measuring in whole blood samples, determination of AR PAS showed very good performance to diagnose sepsis with high sensitivity and specificity, while as a prognostic assay, results suggest potential use to identify good prognosis patients.

Normal AR pathway activity ranges differed between healthy adults and children. This may be “real” differences, for example caused by differences in testosterone levels between adults and children, but lack of differences between men and women make this a less likely explanation. Alternatively, differences may have been caused by different handling of samples between clinical centers involved in pediatric versus adult studies [23],[25],[26],[27],[28],[29],[31],[24].

Since Affymetrix analysis takes too long for the result to be useful in a clinical setting, STP assays have currently been adapted to qPCR for future diagnostic use [15]. The time-to-result of the qPCR-based STP-assays lies within a few hours, and clinical implementation of such assays can be fast, following establishing normal ranges for STP activities. In general, correlation between STP activity determined by Affymetrix-based STP analysis and qPCR-based analysis is very good [14].

Thus, while we show feasibility of determining a normal range of AR pathway activity in whole blood samples based on Affymetrix data analysis, clinical adoption of the AR pathway activity assay for diagnostic use in sepsis will require determination of normal AR pathway activity ranges for both adults and children, using the qPCR-based AR pathway assay, and subsequently further confirmation of diagnostic value.

In addition to diagnosing sepsis, and prediction of prognosis, we hypothesize that the AR pathway activity assay may find clinical utility in prediction of risk at serious infectious complications in the period after surviving sepsis. This is of relevance since patients who survive sepsis often remain immunocompromised for longer periods of time, associated with increased risk at secondary infections with poor clinical outcome [3],[52]. In view of the immunosuppressive role of the AR pathway, we hypothesize that this may be causally related to persistently high AR pathway activity [53].

Similarly, the assay may have value in predicting risk at developing sepsis in patients with a bacterial infection. To illustrate this clinical use case, AR pathway activity was measured in RNAseq data (GSE161731, [54]) from samples of patients presenting either with Community Acquired bacterial Pneumonia (CAP) or with viral influenza infection at the emergency ward, using an RNAseq-converted AR pathway assay (Supplementary Figure S12). In whole blood of CAP patients mean AR pathway activity was increased, compared to healthy individuals or patients with an influenza infection, while around one third of CAP patients had AR pathway activity in the normal range. Within the perspective of the current study, we hypothesize that the latter group with normal AR pathway activity did not have sepsis at admission and was at lower risk to develop sepsis, possibly not needing hospital admission. This is in line with data showing that at least a third of patients presenting with CAP have or develop sepsis [55],[56]. Future clinical studies are necessary to further investigate these clinical use cases.

### STP assays to guide therapy choices

STP activity measurements may be used to develop novel therapies for sepsis, on the premise of a causal relation with sepsis. Targeted drugs are available against the AR, TGFβ, MAPK-AP1, NFκB, and JAK-STAT3 pathways, all involved in immune responses, offering a new perspective on treating sepsis [44]. It is expected that a specific drug will only be effective if the causal signaling pathway(s) is (are) abnormally active. Variation in STP PAS was high in sepsis patients, suggesting that only a subpopulation of patients might benefit from pathway-targeted therapy. Thus, investigation of potential beneficial effects of for example androgen antagonists will require a personalized treatment approach based on measuring respective pathway activity. Interestingly, the AR pathway has been suggested to play a role in severe COVID-19 infections, and an AR inhibitor (proxalutamide) reduced disease severity, overall mortality, and length of hospitalization [57], [58]. Secondary bacterial infections frequently complicate severe COVID-19 pneumonia and we hypothesize that increased AR pathway activity may be associated with immunosuppression and high risk at sepsis, explaining the benefit of anti-androgen therapy in this setting.

### Comparison of our data analysis results with the original data analysis performed in the clinical studies

Since we frequently get questions regarding the difference between our STP analysis and other bioinformatics tools for Pathway Analysis, such as GSEA and Ingenuity, we compared our STP analysis results with data analysis results performed by the investigators who had generated the Affymetrix data. Using a variety of bioanalytical biomarker discovery tools, Ingenuity, PANTHER, D.A.V.I.D., and ToppGene, associations between sepsis and a number of signaling pathways were found that were in agreement with our STP analysis results, that is for the NFκB, MAPK, FOXO-PI3K and JAK-STAT3 pathways, but not for the AR, ER, and TGFβ pathways. Identification of “pathways” using these bioinformatics tools is not suited for analysis of individual sample data and is not informative on signaling pathway activation. Reason that AR, TGFβ, and ER pathways were not identified probably lies in lack of differential expression of signaling genes of these pathways between control and sepsis groups, while we measure differences based on functional pathway activity. To our best knowledge the sepsis-associated gene signatures discovered in these clinical studies have not been implemented in clinical practice.

STP assays were developed for diagnostic purposes and have been analytically validated for measuring signaling pathway activity *prior to use on the current data*. STP results are potentially clinically actionable, since many targeted drugs inhibit activity of the signal transduction pathways that were found to be overactive in sepsis and septic shock.

In summary, our current results show initial results supporting clinical validity and potential clinical utility of measuring activity of the AR and TGFβ signal transduction pathways in patients with sepsis, fulfilling the basic requirements for biomarker assays [59]. In addition, good reproducibility between the different clinical studies suggests that defining a normal (“healthy”) range of pathway activity in whole blood samples will be feasible. This is important for future clinical implementation since usually no individual healthy reference blood sample will be available. We recognize that performing Affymetrix microarrays is not possible in clinical practice. For this reason the STP assays have been converted to qPCR assays, which can be easily performed in a routine hospital lab, and provide a pathway activity score within a few hours, enabling timely clinical decisions [15].

## Supporting information

Supplemental comparison public bioinformatics tools with STP analysis

## Data Availability

Since Affymetrix data to be analyzed in the current study were obtained from the GEO database, and not part of a novel clinical study, trial registration and obtaining informed consent has occurred in the past under the responsibility of the PI (linked to the study-associated publications-referred in the text).

## Limitations and future studies

Clinical studies are needed to confirm the clinical utility of measuring AR (and TGFβ) pathway activity in patients with, or at risk for developing, sepsis or septic shock. The available clinical studies only contained two studies with adult patients. Although results were comparable, causes of sepsis are far more variable in adult patients and in in future studies it would be important to include more adult patient populations.

## Data Availability Statement

All datasets presented in this study are publicly available from the Gene Expression Omnibus (GEO) database [16].

## Author Contributions

WB: Data, results analysis, concept and writing. WV: pathway model development, results analysis. AS: pathway model development, results analysis, and writing. All authors contributed to the article and approved the submitted version.

## Acknowledgments

We acknowledge Yvonne Wesseling-Rozendaal for analyzing dataset GSE161731 and preparing Figure S12; and Paul van de Wiel, Diederick Keizer and Yvonne Wesseling-Rozendaal for critically reviewing the manuscript. We also wish to acknowledge all investigators who generated the GEO datasets used in the current analysis.

