## Supplemental tables and figures for "Androgen receptor pathway activity assay for sepsis diagnosis and prediction of favorable prognosis"

### Supplementary information (I)

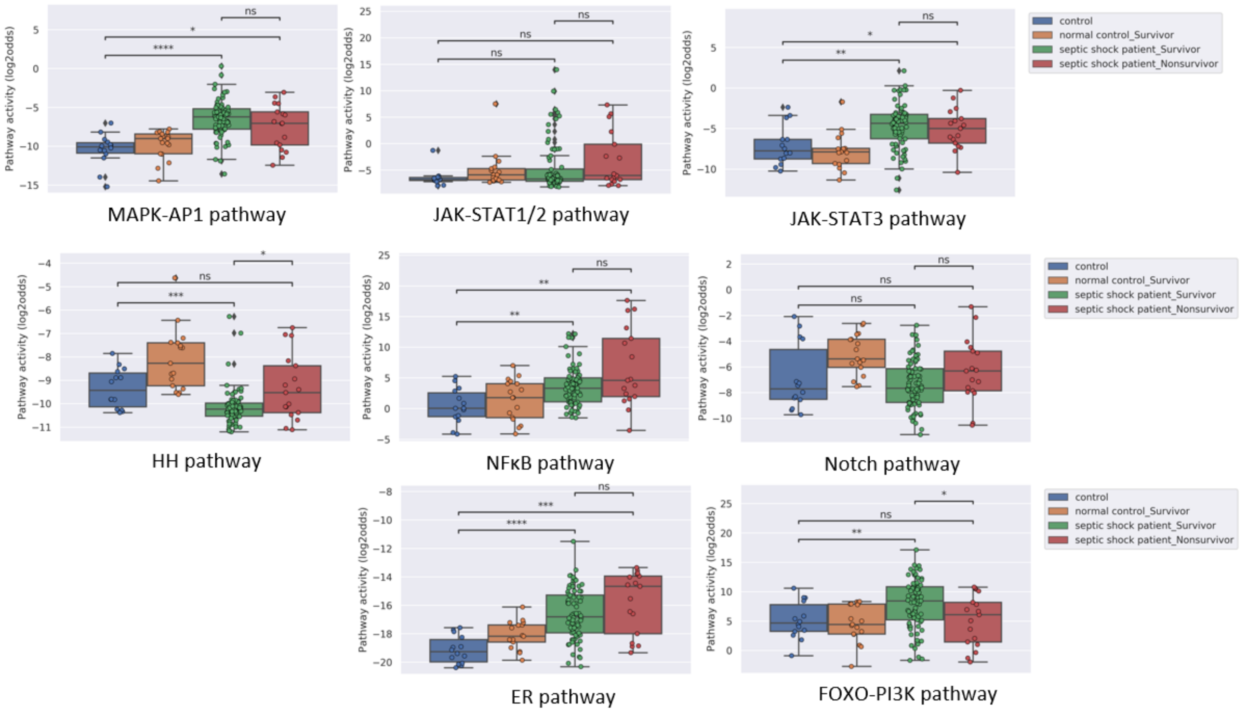


*Figure S1: PAS for pathways NFκB, Notch, ER, FOXO, HH, MAPK-AP1, JAK-STAT3 and JAK-STAT1/2 of dataset GSE26440* [1]. Pathway activity score on Y-axis in log2 odds. Two sided Mann–Whitney–Wilcoxon statistical tests were performed; p-values are indicated in the figures as *p <0.05, **p < 0.01, ***p < 0.001, ****p < 0.0001 or ns (not significant).

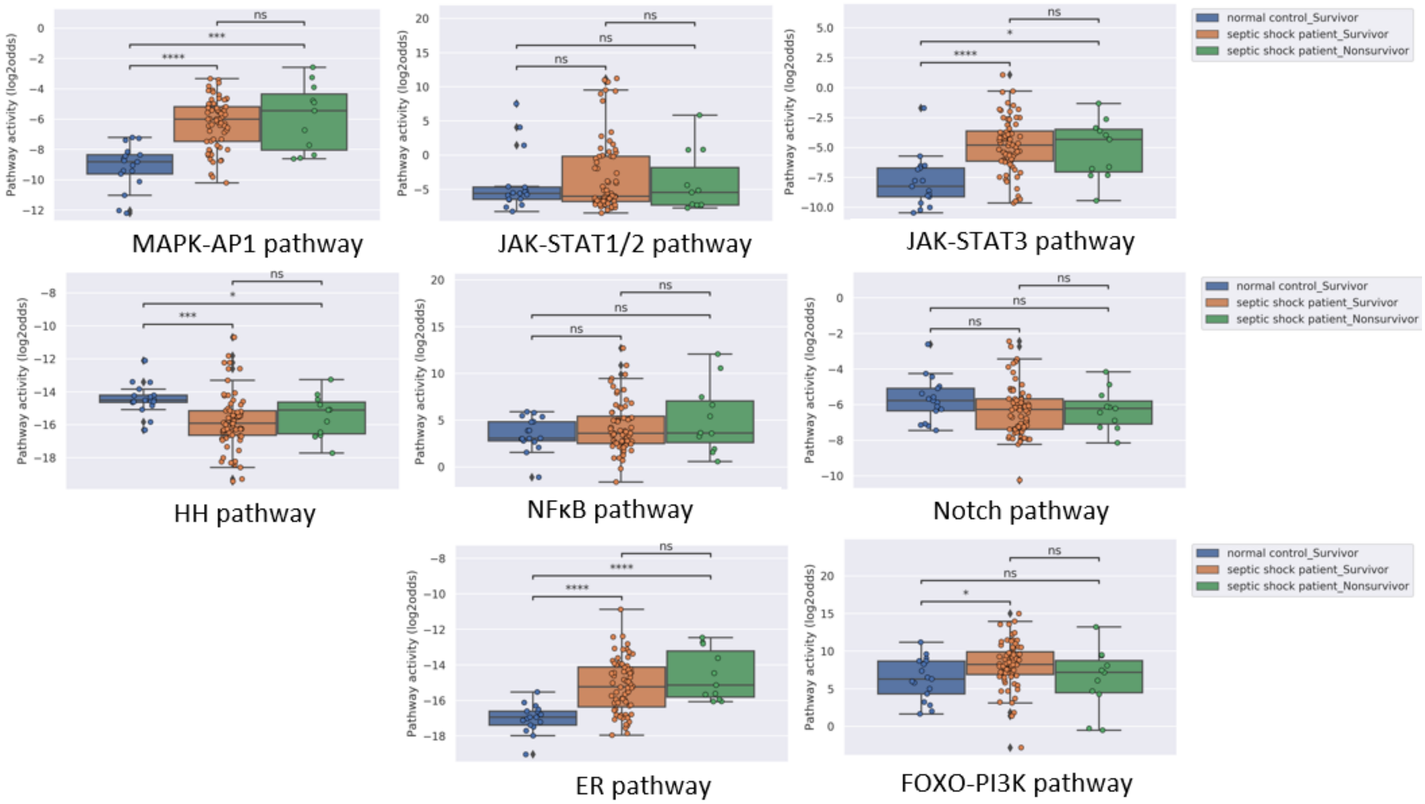


*Figure S2: PAS for pathways NFκB, NOTCH, ER, FOXO-PI3K, HH, MAPK-AP1, JAK-STAT3 and JAK-STAT1/2 of dataset GSE26378* [1]. *Two sided Mann–Whitney–Wilcoxon statistical tests were performed; p-values are indicated in the figures as *p <0.05, **p < 0.01, ***p < 0.001, ****p < 0.0001 or ns (not significant).*

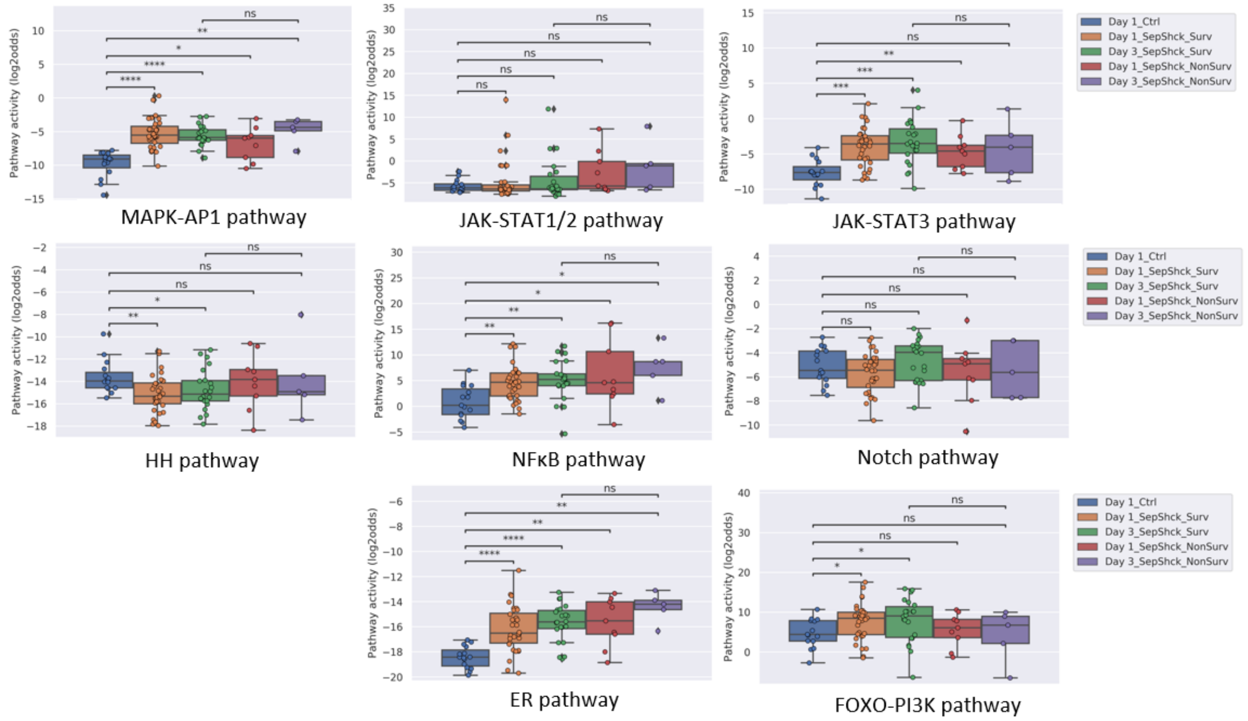


*Figure S3: PAS for pathways NFκKB, Notch, ER, FOXO-PI3K, HH, MAPK-AP1, JAK-STAT3 and JAK-STAT1/2 of dataset GSE4607* [2]. *Two sided Mann–Whitney–Wilcoxon statistical tests were performed; p-values are indicated in the figures as *p <0.05, **p < 0.01, ***p < 0.001, ****p < 0.0001 or ns (not significant).*

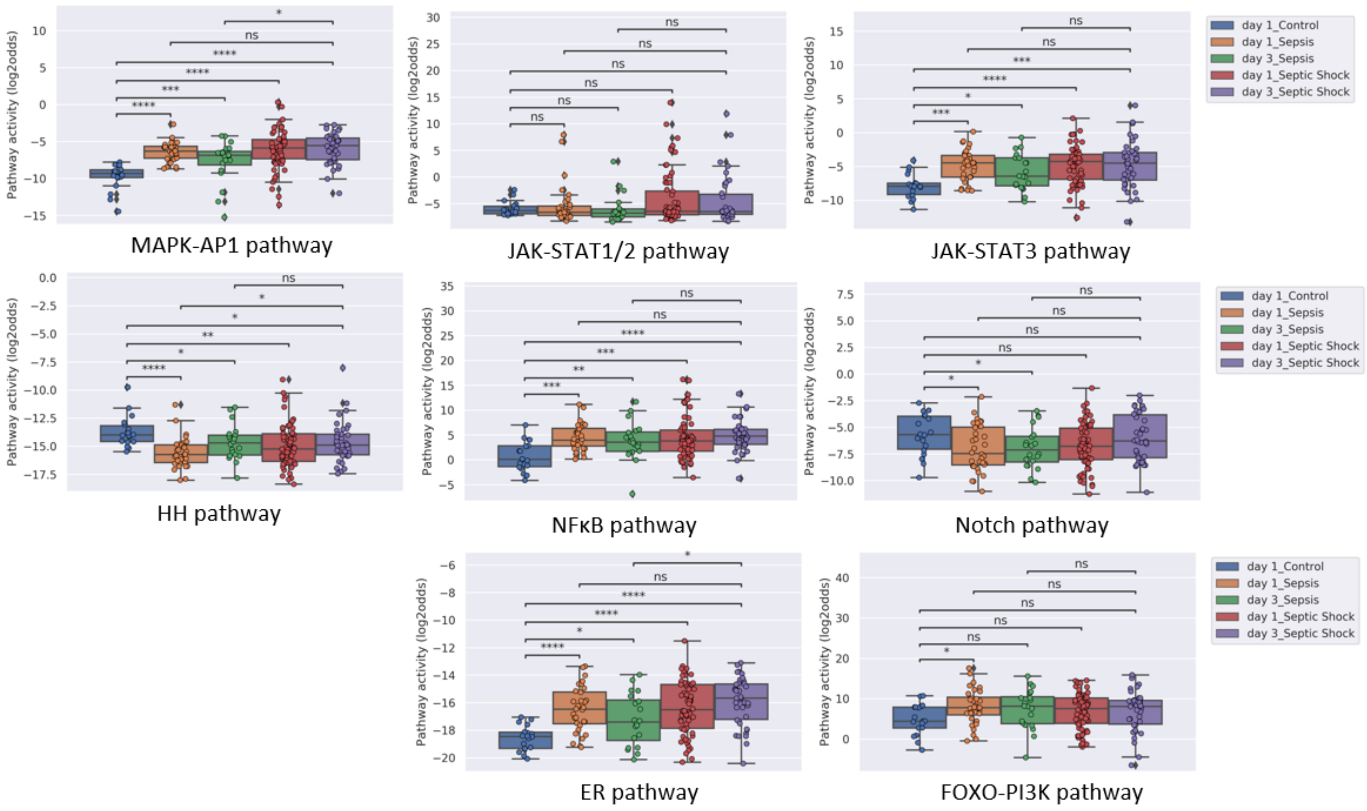


*Figure S4: PAS for pathways NFκB, Notch, ER, FOXO-PI3K, HH, MAPK-AP1, JAK-STAT3 and JAK-STAT1/2 of dataset GSE13904* [3]*. Two sided Mann–Whitney–Wilcoxon statistical tests were performed; p-values are indicated in the figures as *p <0.05, **p < 0.01, ***p < 0.001, ****p < 0.0001 or ns (not significant).*

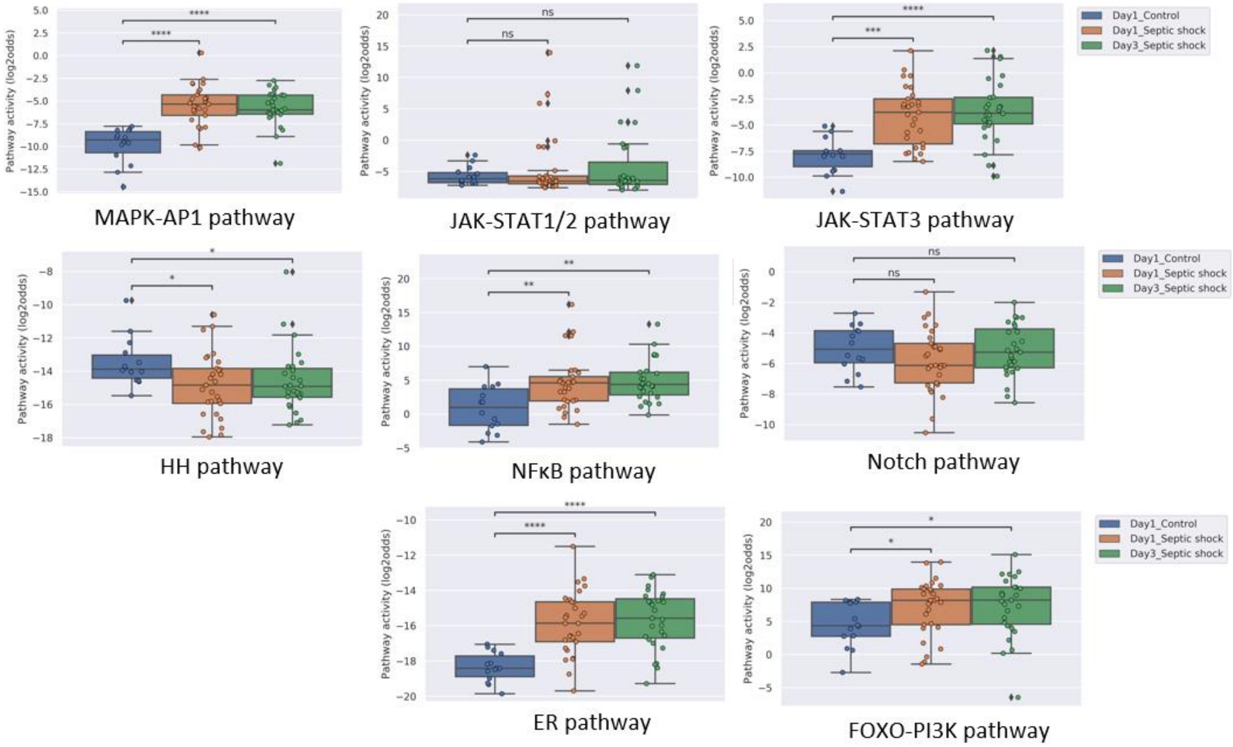


*Figure S5: PAS for pathways NFκB, Notch, ER, FOXO-PI3K, HH, MAPK-AP1, JAK-STAT3 and JAK-STAT1/2 of dataset GSE8121* [4]. *Two sided Mann–Whitney–Wilcoxon statistical tests were performed; p-values are indicated in the figures as *p <0.05, **p < 0.01, ***p < 0.001, ****p < 0.0001 or ns (not significant).*

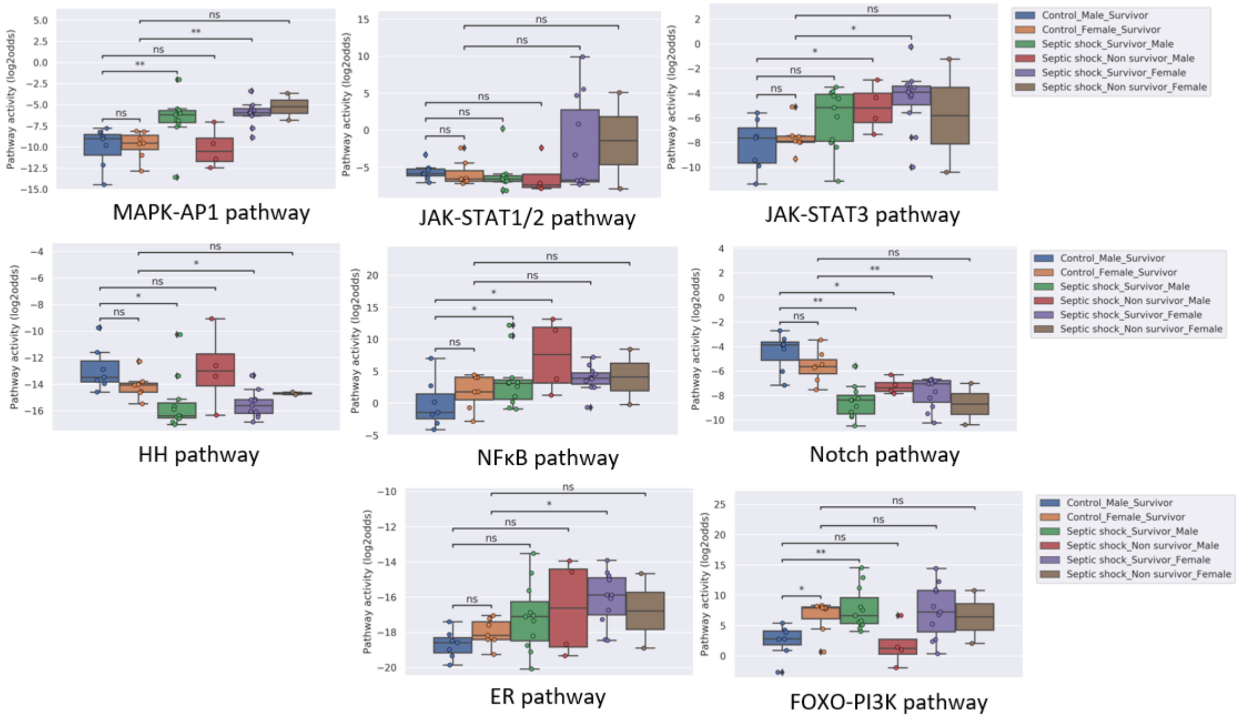


*Figure S6: PAS for pathways NFκB, Notch, ER, FOXO-PI3K, HH, MAPK-AP1, JAK-STAT3 and JAK-STAT1/2 of dataset GSE9692* [5]. *Two sided Mann–Whitney–Wilcoxon statistical tests were performed; p-values are indicated in the figures as *p <0.05, **p < 0.01, ***p < 0.001, ****p < 0.0001 or ns (not significant).*

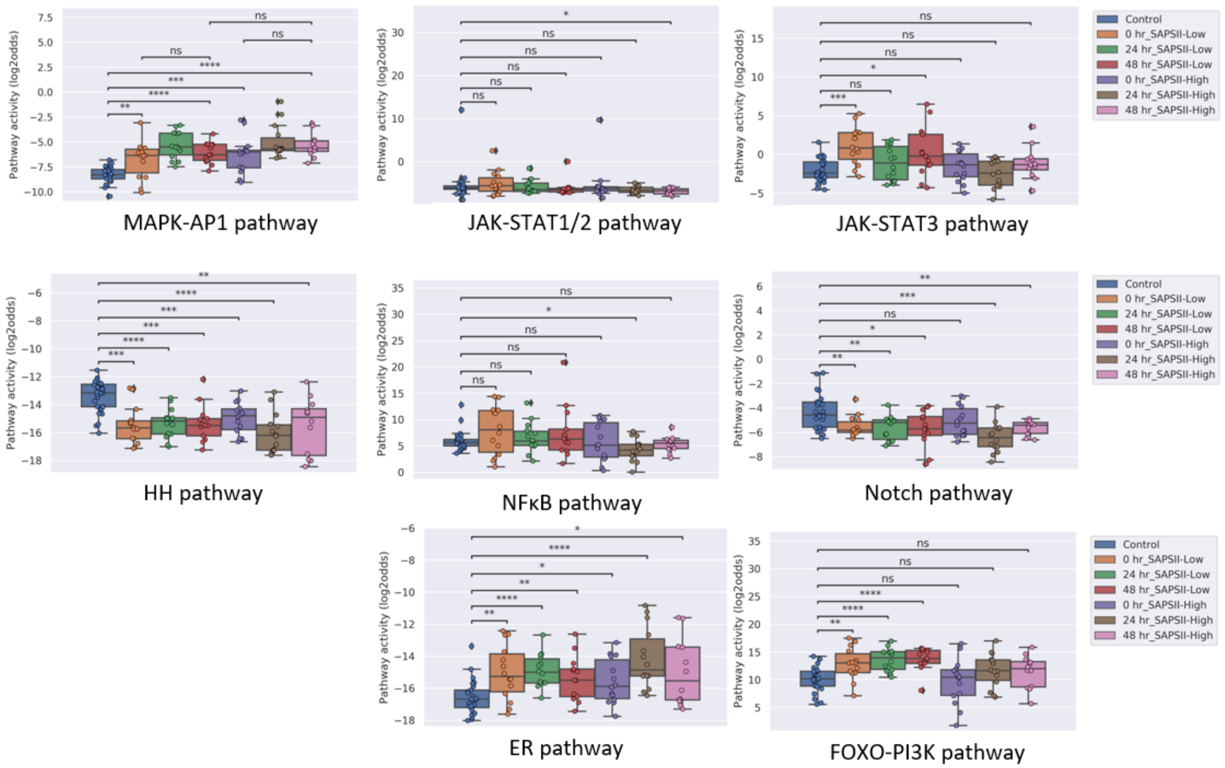


*Figure S7: PAS for pathways NFκB, Notch, ER, FOXO-PI3K, HH, MAPK-AP1, JAK-STAT3 and JAK-STAT1/2 of dataset GSE57065* [6]*. Two sided Mann–Whitney–Wilcoxon statistical tests were performed; p-values are indicated in the figures as *p<0.05, **p < 0.01, ***p < 0.001, ****p < 0.0001 or ns (not significant).*

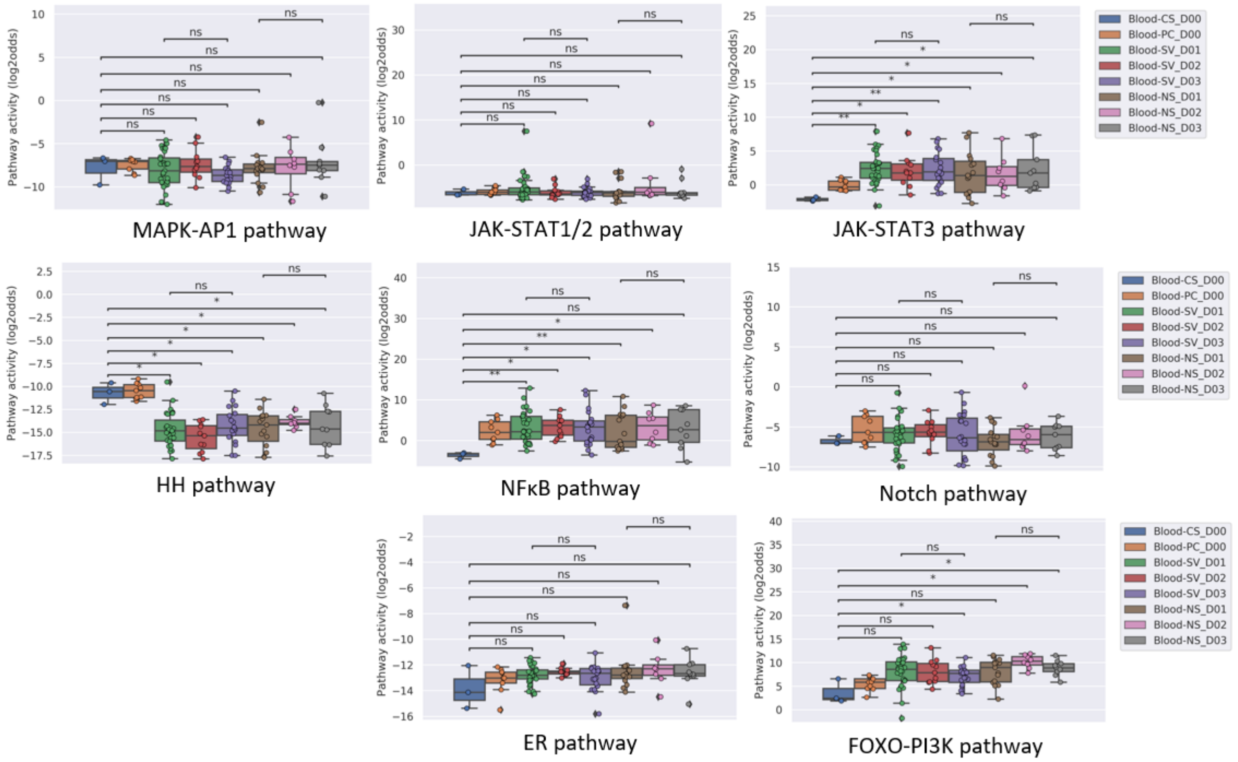


*Figure S8: PAS for pathways NFκB, Notch, , ER, FOXO-PI3K, HH, MAPK-AP1, JAK-STAT3 and JAK-STAT1/2 of dataset GSE95233* [7]. *Two sided Mann–Whitney–Wilcoxon statistical tests were performed; p-values are indicated in the figures as *p <0.05, **p < 0.01, ***p < 0.001, ****p < 0.0001 or ns (not significant).*

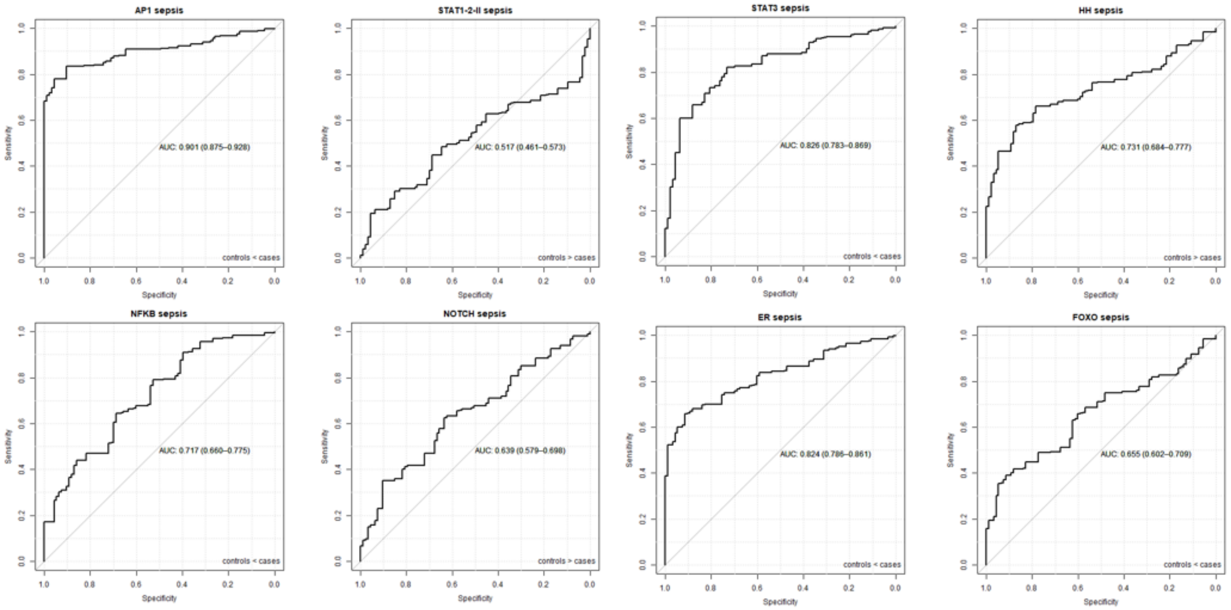


*Figure S9: ROC curves for diagnosis of sepsis for pathways NFκB, Notch, TGFβ, ER, FOXO-PI3K, HH, MAPK-AP1, JAK-STAT3 and JAK-STAT1/2. Controls and sepsis patients (sepsis and sepsis shock) from dataset* GSE26440 [1], GSE26378 [1], GSE4607 [2], GSE13904 [3], GSE8121[4] and GSE9692 [5] *are included.*

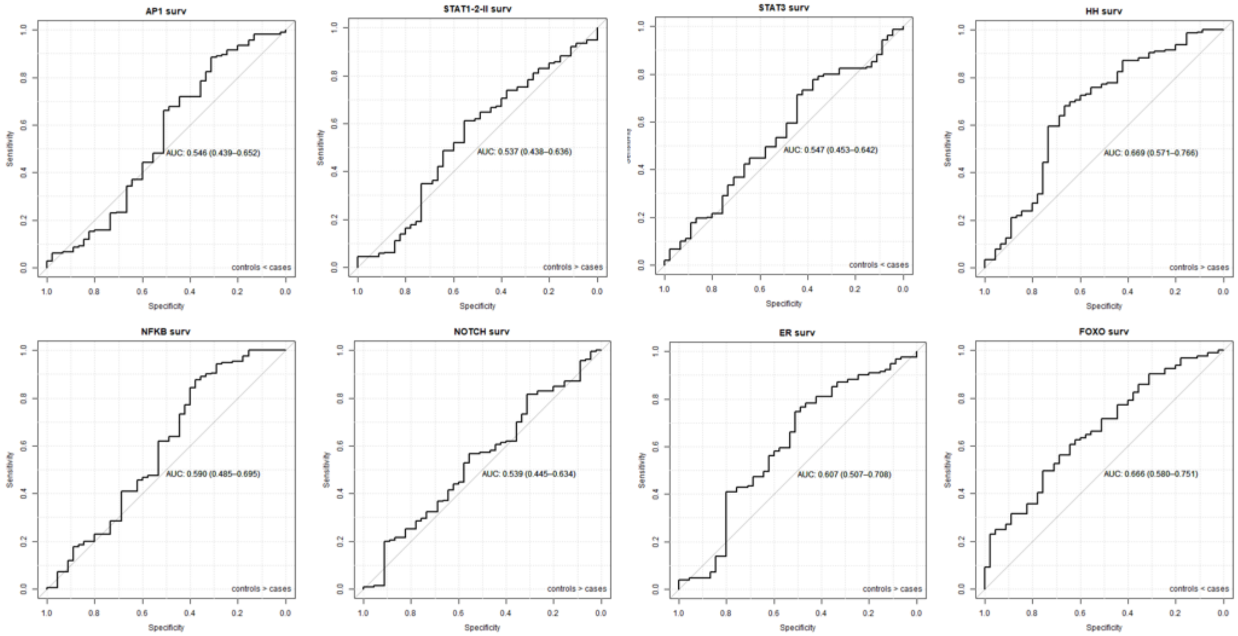


*Figure S10: ROC curves for survival of sepsis for pathways NκKB, Notch, TGFβ, ER, FOXO-PI3K, HH, MAPK-AP1, JAK-STAT3 and JAK-STAT1/2 sepsis survival and non survival patients from dataset GSE26440* [1]*, GSE26378* [1]*, GSE4607* [2]*, and GSE9692* [5] *are included.*

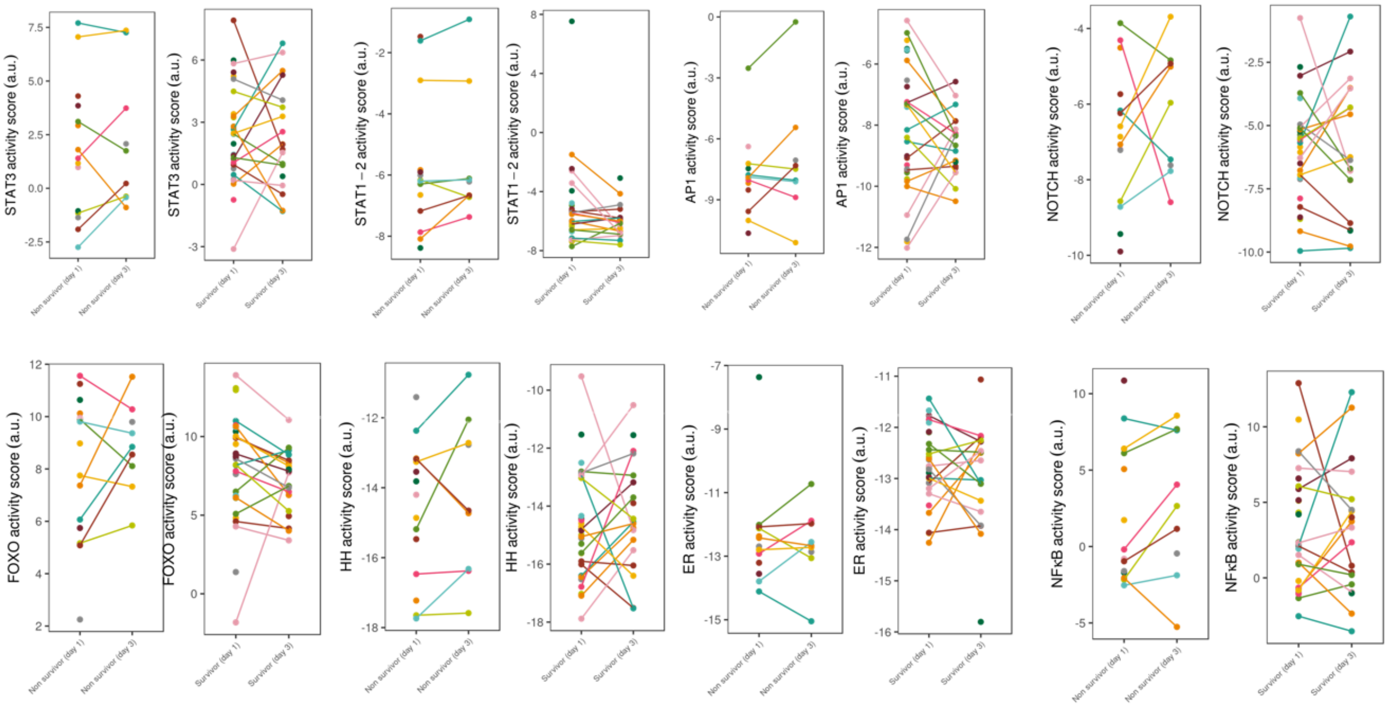


*Figure S11: PAS of day 1 versus day 3 of sepsis survival and non-survival patients for pathways NFκB, Notch, TGFβ, ER, FOXO-PI3K, HH, MAPK-AP1, JAK-STAT3 and JAK-STAT1/2. Dataset GSE95233* [7].

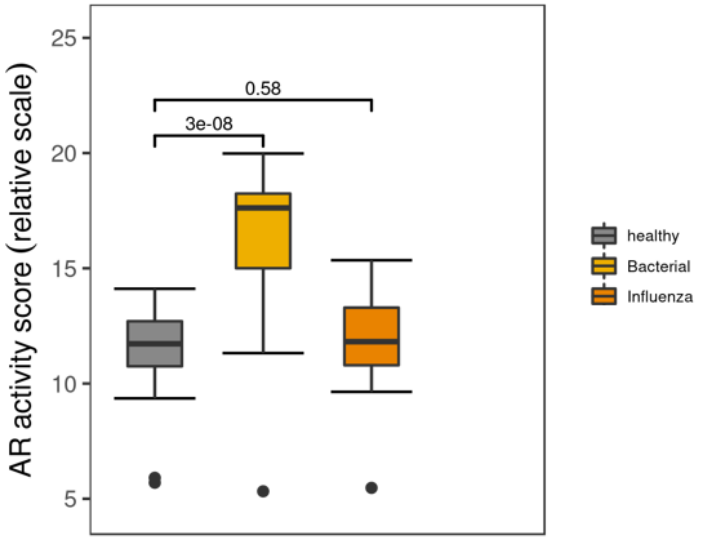


Figure S12: AR pathway activity analysis of dataset GSE161731 [8] of whole blood samples from patients presenting to the emergency department with acute respiratory infection caused by influenza (virus), or bacterial pneumonia and matched healthy controls. RNAseq data were analyzed with our signal transduction pathway assays converted to RNAseq as input data. *Two sided Mann–Whitney–Wilcoxon statistical tests is performed; p-values are indicated in the figures.*

| Dataset | Sample type | Sample prep | Population information | GSM numbers of removed duplicates | Pubmed |
|---|---|---|---|---|---|
| GSE26440 | blood samples were obtained within 24 hours of initial presentation to the PICU with septic shock | Whole blood and PaxGene™ Blood RNA System | Children =10 years of age admitted to the pediatric intensive care unit (PICU) and meeting pediatric-specific criteria for septic shock were eligible for enrollment | GSM648594<br>GSM648650<br>GSM648652<br>GSM648653<br>GSM648657<br>GSM648675<br>GSM648680<br>GSM648719 | https://pubmed.ncb |
| GSE26378 | blood samples were obtained within 24 hours of initial presentation to the PICU with septic shock | Whole blood and PaxGene™ Blood RNA System | Children =10 years of age admitted to the pediatric intensive care unit (PICU) and meeting pediatric-specific criteria for septic shock were eligible for enrollment | GSM647621 | https://pubmed.ncbi.[illegible] |
| GSE4607 | Blood samples were obtained within 24 hours of admission to the PICU, heretofore referred to as “Day 1” of septic shock. | Whole blood and PaxGene™ Blood RNA System | Children < 10 years of age admitted to the pediatric intensive care unit (PICU) and meeting criteria for septic shock were eligible for the study. Septic shock was defined using pediatric-specific criteria | | https://pubmed.ncb |
| GSE13904 | blood samples were obtained on day 1 of the study and when possible on day 3 of the study. Day 1 of the study was defined as within the first 24 hours of meeting criteria for a given study category in any PICU patient, whether at admission to the PICU or after initial admission to the PICU with a nonstudy-related classification. Day 3 of the study was defined as 48 hours after drawing day 1 samples, regardless of the PICU status. | Whole blood and PaxGene™ Blood RNA System | Children ≤10 years of age admitted to the pediatric intensive care units (PICUs) and meeting published, pediatric-specific criteria for SIRS, sepsis, or septic shock were eligible | GSM350141<br>GSM350142<br>GSM350144<br>GSM350145<br>GSM350146<br>GSM350147<br>GSM350154<br>GSM350209<br>GSM350238<br>GSM350239<br>GSM350246<br>GSM350248<br>GSM350249<br>GSM350257<br>GSM350262<br>GSM350267<br>GSM350268<br>GSM350271<br>GSM350272<br>GSM350273<br>GSM350274<br>GSM350275<br>GSM350279 | https://pubmed.ncb |

| | | | | | |
|---|---|---|---|---|---|
| | | | | GSM350286<br>GSM350291<br>GSM350293<br>GSM350294<br>GSM350297<br>GSM350298<br>GSM350299<br>GSM350300<br>GSM350302<br>GSM350303<br>GSM350304<br>GSM350309<br>GSM350310<br>GSM350316<br>GSM350317<br>GSM350325<br>GSM350326<br>GSM350343<br>GSM350344<br>GSM350347<br>GSM350348<br>GSM350350<br>GSM350351<br>GSM350358<br>GSM350363 | |
| GSE8121 | Blood samples for RNA isolation were obtained within 24 h of admission to the PICU, heretofore called "day one" of septic shock. An additional sample was obtained for RNA isolation 48 hours after obtaining the day one sample ("day three" of septic shock). | Whole blood and PaxGene™ Blood RNA System | Children < ten years of age admitted to the PICU and meeting criteria for septic shock were eligible for the study | GSM201231<br>GSM201241<br>GSM201242<br>GSM201245<br>GSM201246<br>GSM201251<br>GSM201252<br>GSM201254<br>GSM201256<br>GSM201259<br>GSM201261<br>GSM201262<br>GSM201263<br>GSM201265<br>GSM201268<br>GSM201270<br>GSM201272<br>GSM201273<br>GSM201281<br>GSM201282 | https://pubmed.ncb |

| | | | | | |
|---|---|---|---|---|---|
| | | | | GSM201283<br>GSM201286<br>GSM201291<br>GSM201292<br>GSM201295<br>GSM201297<br>GSM201299<br>GSM201301 | |
| GSE9692 | blood samples for RNA isolation were obtained within 24 hours of admission to the PICU, heretofore referred to as “day 1” of septic shock. | Whole blood and PaxGene™ Blood RNA System | Children < 10 years of age admitted to the pediatric intensive care unit (PICU), and meeting pediatric-specific criteria for septic shock were eligible for the study | GSM244479<br>GSM244483<br>GSM244484<br>GSM244486<br>GSM244490<br>GSM244491<br>GSM244492<br>GSM244497<br>GSM244504<br>GSM244507 | https://pubmed.ncb |
| GSE57065 | Blood samples were collected within 30 min and 24 and 48 h after shock. | Whole blood and PaxGene™ Blood RNA System | 18 years and above were enrolled at the onset of septic shock. | | https://pubmed.ncb |
| GSE95233 | Septic shock patients were sampled twice, at admission, and a second time at D2 or D3. | Whole blood and PaxGene™ Blood RNA System | Septic shock patients (adults) admitted to the Intensive Care Unit (ICU) were compared to healthy volunteers, and according to day 28 survival status. | | https://pubmed.ncb |
| GSE161731 | whole blood samples from patients presenting to the emergency department with acute respiratory infection caused by influenza (virus), or bacterial pneumonia and matched healthy controls | Whole blood and PaxGene™ Blood RNA System | Patients presenting to the emergency department with acute respiratory infections | | https://pubmed.ncb |

**Supplementary Table 1**: Public clinical datasets from the GEO database used in used in study.

| | Healthy controls Children (log2 odds scores) | | | |
|---|---|---|---|---|
| **Pathway** | **Mean** | **SD** | **Mean + 1 SD** | **Mean + 2 SD** |
| MAPK-AP1 | -9.67 | 1.73 | -7.94 | -6.20 |
| AR | -15.37 | 1.52 | -13.85 | -12.33 |
| ER | -18.22 | 1.10 | -17.12 | -16.02 |
| HH* | -13.86 | 1.45 | -15.31 | -16.76 |
| NFκB | 1.49 | 3.01 | 4.50 | 7.51 |
| Notch | -5.61 | 1.78 | -3.84 | -2.06 |
| FOXO-PI3K | 5.12 | 3.33 | 8.45 | 11.77 |
| JAK-STAT1/2 | -5.54 | 2.34 | -3.20 | -0.86 |
| JAK-STAT3 | -7.86 | 2.00 | -5.86 | -3.85 |
| TGFβ | -17.05 | 1.63 | -15.42 | -13.79 |

| | Healthy controls Adults (log2 odds scores) | | | |
|---|---|---|---|---|
| **Pathway** | **Mean** | **SD** | **Mean + 1 SD** | **Mean + 2 SD** |
| MAPK-AP1 | -8.04 | 0.94 | -7.10 | -6.16 |
| AR | -12.44 | 1.44 | -11.01 | -9.57 |
| ER | -15.53 | 1.88 | -13.65 | -11.77 |
| HH* | -12.51 | 1.73 | -10.78 | -9.04 |
| NFκB | 4.29 | 3.50 | 7.79 | 11.29 |
| Notch | -4.74 | 1.62 | -3.12 | -1.50 |
| FOXO-PI3K | 8.49 | 3.22 | 11.70 | 14.92 |
| JAK-STAT1/2 | -5.59 | 3.12 | -2.47 | 0.65 |
| JAK-STAT3 | -1.61 | 1.54 | -0.07 | 1.47 |
| TGFβ | -13.01 | 1.83 | -11.18 | -9.35 |

| | Healthy controls Children + Adults (log2 odds scores) | | | |
|---|---|---|---|---|
| **Pathway** | **Mean** | **SD** | **Mean + 1 SD** | **Mean + 2 SD** |
| MAPK-AP1 | -9.21 | 1.71 | -7.49 | -5.78 |
| AR | -14.54 | 2.00 | -12.54 | -10.54 |
| ER | -17.46 | 1.83 | -15.63 | -13.80 |
| HH* | -13.48 | 1.65 | -15.13 | -16.77 |
| NFκB | 2.29 | 3.39 | 5.68 | 9.06 |
| Notch | -5.36 | 1.77 | -3.59 | -1.82 |

| | | | | |
|---|---|---|---|---|
| FOXO-PI3K | 6.08 | 3.62 | 9.70 | 13.32 |
| JAK-STAT1/2 | -5.56 | 2.57 | -2.98 | -0.41 |
| JAK-STAT3 | -6.08 | 3.40 | -2.69 | 0.71 |
| TGFβ | -15.90 | 2.49 | -13.41 | -10.92 |

**Supplementary Table 2**: Mean plus 1 and 2 SD of pathway activity scores in healthy children (n=93; GSE26440, GSE26378, GSE4607, GSE9692, GSE8121 and GSE13904) and adults (n=37; GSE57065 and GSE95233). *for HH mean minus SD.

| T-test one sided | Survivors day 1 vs day 3 | | | Non survivors day 1 vs day 3 | | |
|---|---|---|---|---|---|---|
| | paired | | | paired | | |
| | p-value | significance | numbers day 1 / day 3 | p-value | significance | numbers day 1 / day 3 |
| MAPK-AP1 | 0.37 | - | 17/17 | 0.82 | - | 8/8 |
| AR | 0.19 | - | 17/17 | 0.06 | - | 8/8 |
| ER | 0.36 | - | 17/17 | 0.73 | - | 8/8 |
| FOXO-PI3K | 0.23 | - | 17/17 | 0.85 | - | 8/8 |
| HH | 0.67 | - | 17/17 | 0.79 | - | 8/8 |
| NFκB | 0.57 | - | 17/17 | 0.92 | - | 8/8 |
| Notch | 0.57 | - | 17/17 | 0.67 | - | 8/8 |
| JAK-STAT1/2 | 0.06 | - | 17/17 | 0.93 | - | 8/8 |
| JAK-STAT3 | 0.58 | - | 17/17 | 0.73 | - | 8/8 |
| TGFβ | 0.40 | - | 17/17 | 0.61 | - | 8/8 |

| T-test one sided | Survivors day 1 vs day 3 | | | Non survivors day 1 vs day 3 | | |
|---|---|---|---|---|---|---|
| | unpaired | | | unpaired | | |
| | p-value | significance | numbers day 1 / day 3 | p-value | significance | numbers day 1 / day 3 |
| MAPK-AP1 | 0.12 | - | 31/19 | 0.74 | - | 15/9 |
| AR | 0.03 | * | 31/19 | 0.31 | - | 15/9 |
| ER | 0.24 | - | 31/19 | 0.41 | - | 15/9 |
| FOXO-PI3K | 0.11 | - | 31/19 | 0.79 | - | 15/9 |
| HH | 0.79 | - | 31/19 | 0.67 | - | 15/9 |
| NFκB | 0.43 | - | 31/19 | 0.60 | - | 15/9 |
| Notch | 0.50 | - | 31/19 | 0.79 | - | 15/9 |
| JAK-STAT1/2 | 0.09 | - | 31/19 | 0.62 | - | 15/9 |
| JAK-STAT3 | 0.44 | - | 31/19 | 0.66 | - | 15/9 |
| TGFβ | 0.05 | - | 31/19 | 0.49 | - | 15/9 |

**Supplementary Table 3:** Paired and unpaired one sided t-test for survivors and non survivors day 1 vs day 3 from dataset GSE95233. **p <0.05, **p < 0.01, - not significant.*

| | Mean | | | | | | | |
|---|---|---|---|---|---|---|---|---|
| | Children | | | | | | Adults | |
| Pathway | GSE4607 | GSE26440 | GSE8121 | GSE9692 | GSE13904 | GSE26378 | GSE57065 | GSE95233 |
| MAPK-AP1 | -9.8 | -10.1 | -9.6 | -9.3 | -9.5 | -9.1 | -8.3 | -7.5 |
| AR | -15.3 | -15.2 | -15.3 | -15.3 | -15.7 | -15.6 | -13.1 | -11.0 |
| ER | -18.4 | -18.6 | -18.4 | -18.0 | -18.7 | -17.0 | -16.6 | -13.3 |
| HH | -13.6 | -14.2 | -13.4 | -13.3 | -13.6 | -14.4 | -13.4 | -10.6 |
| NFκB | 0.8 | 1.0 | 1.0 | 2.0 | 1.0 | 3.4 | 6.0 | 0.8 |
| Notch | -5.1 | -5.9 | -5.1 | -5.0 | -6.2 | -5.9 | -4.3 | -5.7 |
| FOXO-PI3K | 4.9 | 4.9 | 4.6 | 5.1 | 5.1 | 6.1 | 10.2 | 5.0 |
| JAK-STAT1/2 | -5.7 | -5.6 | -5.5 | -5.6 | -5.8 | -5.0 | -5.4 | -5.9 |
| JAK-STAT3 | -7.7 | -7.5 | -8.1 | -8.2 | -8.1 | -8.3 | -2.1 | -0.6 |
| TGFβ | -17.0 | -17.4 | -17.1 | -16.6 | -16.8 | -16.8 | -13.2 | -12.6 |

| | SD | | | | | | | |
|---|---|---|---|---|---|---|---|---|
| | Children | | | | | | Adults | |
| Pathway | GSE4607 | GSE26440 | GSE8121 | GSE9692 | GSE13904 | GSE26378 | GSE57065 | GSE95233 |
| MAPK-AP1 | 1.9 | 2.0 | 1.7 | 1.5 | 1.2 | 1.5 | 0.8 | 1.0 |
| AR | 1.4 | 1.8 | 1.2 | 1.3 | 1.3 | 1.7 | 1.2 | 0.6 |
| ER | 0.9 | 1.1 | 0.9 | 0.9 | 1.0 | 0.8 | 1.1 | 1.2 |
| HH | 1.5 | 1.4 | 1.6 | 1.8 | 1.6 | 1.0 | 1.2 | 0.9 |
| NFκB | 3.2 | 3.1 | 3.3 | 3.3 | 2.5 | 1.8 | 1.9 | 3.5 |
| Notch | 2.0 | 1.6 | 1.5 | 1.6 | 2.0 | 1.5 | 1.4 | 1.6 |
| FOXO-PI3K | 1.1 | 2.6 | 1.2 | 1.3 | 1.3 | 2.4 | 2.2 | 1.9 |
| JAK-STAT1/2 | 1.9 | 2.3 | 1.8 | 2.0 | 2.1 | 1.5 | 3.8 | 0.7 |
| JAK-STAT3 | 1.6 | 1.6 | 1.4 | 1.0 | 1.3 | 2.3 | 1.5 | 1.2 |
| TGFβ | 3.0 | 3.1 | 2.3 | 2.2 | 3.1 | 3.1 | 2.1 | 1.2 |

**Supplementary Table 4:** Mean and standard deviation PAS of each dataset including healthy children and adults.

| Sepsis diagnosed | sepsis confirmed | No sepsis | |
|---|---|---|---|
| Test positive | 308 | 3 | 311 |
| Test negative | 91 | 90 | 181 |
| | 399 | 93 | 492 |

| | |
|---|---|
| Sensitivity | 77% |
| Specificity | 97% |
| Possitive prediction value (PPV) | 99% |
| Negative prediction value (NPV) | 50% |
| Fisher's Exact Test | p-value < 2.2e-16 |

| Dying from sepsis | sepsis nonsurvival | sepsis survival | |
|---|---|---|---|
| Test positive | 42 | 157 | 199 |
| Test negative | 3 | 53 | 56 |
| | 45 | 210 | 255 |

| | |
|---|---|
| Sensitivity | 93% |
| Specificity | 25% |
| Possitive prediction value (PPV) | 21% |
| Negative prediction value (NPV) | 95% |
| Fisher's Exact Test | p-value = 0.005023 |

| Surviving sepsis | sepsis survival | sepsis non survival | |
|---|---|---|---|
| Test positive (below threshold) | 53 | 3 | 56 |
| Test negative (above threshold) | 157 | 42 | 199 |
| | 210 | 45 | 255 |

| | |
|---|---|
| Sensitivity | 25% |
| Specificity | 93% |
| Possitive prediction value (PPV) | 95% |
| Negative prediction value (NPV) | 21% |
| Fisher's Exact Test | p-value = 0.005023 |

**Supplementary table 6: TGFβ sensitivity, specificity, PPV and NPV.**

| Sepsis diagnosed | sepsis confirmed | No sepsis | |
|---|---|---|---|
| Test positive | 254 | 2 | 256 |
| Test negative | 145 | 91 | 236 |
| | 399 | 93 | 492 |

| | |
|---|---|
| Sensitivity | 64% |
| Specificity | 98% |
| Possitive prediction value (PPV) | 99% |
| Negative prediction value (NPV) | 39% |
| Fisher's Exact Test | p-value < 2.2e-16 |

| Dying from sepsis | sepsis nonsurvival | sepsis survival | |
|---|---|---|---|
| Test positive | 31 | 142 | 173 |
| Test negative | 14 | 68 | 82 |
| | 45 | 210 | 255 |

| | |
|---|---|
| Sensitivity | 69% |
| Specificity | 32% |
| Possitive prediction value (PPV) | 18% |
| Negative prediction value (NPV) | 83% |

Fisher's Exact Test p-value = 1

| Surviving sepsis | sepsis survival | sepsis non survival | |
|---|---|---|---|
| Test positive (below threshold) | 68 | 14 | 82 |
| Test negative (above threshold) | 142 | 31 | 173 |
| | 210 | 45 | 255 |

| | |
|---|---|
| Sensitivity | 32% |
| Specificity | 69% |
| Possitive prediction value (PPV) | 83% |
| Negative prediction value (NPV) | 18% |
| Fisher's Exact Test | p-value = 1 |

**Supplementary table 5: AR pathway assay for sepsis diagnosis and prognosis: sensitivity, specificity, PPV and NPV.**
