## Supplemental comparison public bioinformatics tools with STP analysis for "Androgen receptor pathway activity assay for sepsis diagnosis and prediction of favorable prognosis"

### Supplementary information (II)

**Comparison of clinical study data analysis results described in previous publications coupled to the Affymetrix datasets with results obtained with STP (Signal Transduction Pathway) analysis.**

For each publication in which an Affymetrix dataset had been analyzed that was also analyzed for signaling pathway activity in the current study, we have listed the used bioinformatics tools for data analysis, together with obtained results with respect to functional gene annotations and identified "pathways" as defined by the used tools (Ingenuity, PANTHER, D.A.V.I.D., ToppGene), and finally the data-analysis-related conclusions taken from the associated publication abstract.

Comparing the "pathway" results with the results of the STP analysis described in the current paper illustrates the differences between the two approaches (as described before,[1], [2]). First, there is hardly or no overlap between the “pathways” identified by the use of Ingenuity/PANTHER/DAVID/ToppGene, and the signal transduction pathways that were shown in the current analysis to be abnormally active in patients with sepsis/septic shock, and related to survival. The PR, AR, ER, and TGFβ pathways were not mentioned in any of the previous analyses results. MAPK and NFkB pathways came up a few times as linked to sepsis, though their activation state remained unknown. IL6 and IL10 pathway signaling came up, which can be linked to the JAK-STAT3 pathway signaling pathway, however, no information on the activation state of the pathway was provided by the analysis. Mostly differentially expressed genes were be linked to all kinds of physiological mechanisms, by means of functional annotation, or to immune processes like T cell or B cell activation, dendritic cell maturation, etc (see below). Though conceivable that such cellular processes may be more prominent in patients with sepsis, and lead to new hypothesis regarding the pathophysiology, their description is vague. Identification does not directly lead to new therapeutic approaches, since these cellular processes do not present targets for existing drugs/treatments, and therefore are not clinically actionable. Also the effect of the drug on such processes cannot be measured, especially not in a quantitative manner. Similarly, they may provide leads to develop diagnostic/prognostic tests, however there remains a long track for validation of any of the gene classifiers. This includes development of an mRNA-based assay which is applicable to a single sample (as is the case for the STP assays).

Sofar, to the best of our knowledge, none of such classifiers made it to clinical implementation.

As an example, while a carefully validated (on an independent dataset) relation between low Zn levels and sepsis was found ([3]), the clinical relevance of zinc levels for prognosis of sepsis patients has subsequently not been confirmed in clinical studies and current guidelines for pediatric sepsis treatments advise against zinc administration in view of lack of clinical evidence [4]. Thus, while bioinformatics analysis revealed a robust *association* between decreased Zn levels and sepsis, a causal relation was probably lacking. This resulted in a negative clinical study and negative advice with respect to Zn supplementation.

In summary, bioinformatics tools such as Ingenuity, define a "pathway" as "any cellular (biochemical) process", and have been developed for gene expression-based *biomarker discovery purposes* by comparing groups of sample data, and as such, for hypothesis generation. Development of diagnostic assays or identification of a new drug target can start from there.

**Summary identified signaling pathways (expression of signaling pathway components, without indication about pathway activity status):**

1. *NFκB / TLR (associated with NFκB pathway):* [5],[6],[3],[7]
2. *P38-MAPK /PDGF (associated with MAPK-AP1 pathway):* [6],[3],[8],[7]
3. *IL6/IL10 (associated with JAK-STAT3 pathway):* [6],[3],[8],[7]
4. *Integrin signaling/Insulin signaling/IGF1 signaling (associated with PI3K pathway):* [6]
5. *Wnt (associated with the Wnt pathway):* [6]

**<u>Description of bioinformatics tools used and obtained results per clinical study</u>**

**J. L. Wynn *et al.*, “The influence of developmental age on the early transcriptomic response of children with septic shock,” *Mol Med*, vol. 17, no. 11–12, pp. 1146–1156, 2011, doi: 10.2119/molmed.2011.00169. Dataset GSE26440 (GSE26378)**

*Bioinformatics tools used to identify cellular pathways (not specifically signal transduction pathways)*

- Ingenuity

*Identified “pathways” in pediatric sepsis patients:*

1. B-cell receptor signaling
2. TREM1 signaling
3. Pattern recognition receptor signaling
4. NF-κB signaling
5. Dendritic cell maturation
6. Communication between innate and adaptive immunity

*Conclusions taken from publication-abstract:*

- Sepsis: reduced expression of genes representing key pathways of innate and adaptive immunity
- Sepsis: predominantly downregulated transcriptome
- Sepsis: Neonates and school-age subjects had the most uniquely regulated genes relative to controls.

General: “Age-specific studies of the host response are necessary to identify developmentally relevant translational opportunities that may lead to improved sepsis outcomes.”

**H. R. Wong *et al.*, “Genome-level expression profiles in pediatric septic shock indicate a role for altered zinc homeostasis in poor outcome,” *Physiol Genomics*, vol. 30, no. 2, pp. 146–155, Jul. 2007, doi: 10.1152/physiolgenomics.00024.2007. Dataset GSE4607**

*Bioinformatics tools used to identify cellular pathways (not specifically signal transduction*

- D.A.V.I.D. (Database for Annotation, Visualization and Integrated Discovery
- Ingenuity Pathway Analysis, limited to “canonical pathways”

*Results data analysis*

*D.A.V.I.D.: top functional gene annotations, upregulated genes:*

1. direct protein sequencing
2. response to other organism
3. response to pest/pathogen/parasite
4. glycoprotein
5. response to stress
6. membrane
7. response to biotic stimulus
8. response to wounding
9. signal

10. inflammatory response
11. response to external stimulus
12. defense response
13. immune response
14. protein binding
15. lipoprotein
16. phosphorylation
17. intracellular signaling cascade
18. plasma membrane
19. signal transduction
20. protein kinase cascade

*D.A.V.I.D.: Top 20 functional annotations, downregulated genes in patients with septic shock*

1. nuclear protein
2. zinc
3. zinc-finger
4. transcription
5. nucleus
6. transcription regulation
7. dna-binding
8. metal-binding
9. t-cell
10. nucleic acid binding

11. zinc ion binding

12. membrane-bound organelle

13. regulation of intracellular physiological process

14. regulation of cellular process

15. defense response

16. nucleobase, nucleoside, nucleotide, and nucleic acid metabolism

17. regulation of physiological process

18. immune response

19. antigen processing/presentation

20. regulation of biological process

*Ingenuity Pathways analysis (canonical): upregulated genes*

1. Interleukin-6 signaling
2. Interleukin-10 signaling
3. Toll-like receptor signaling
4. B-cell receptor signaling
5. Integrin signaling
6. Complement and coagulation cascades
7. Granulocyte/macrophage-colony stimulation factor signaling
8. p38 MAP kinase signaling
9. Leukocyte extravasation signaling
10. NF-κB signaling

Ingenuity Pathways Analysis (canonical), downregulated genes:

1. T-cell receptor signaling
2. Antigen presentation pathway
3. Natural killer cell signaling
4. Cell Cycle: G1/S checkpoint regulation
5. N-glycan biosynthesis

*Conclusions taken from publication-abstract:*

- genome-level alterations of zinc homeostasis

**H. R. Wong *et al.*, "Genomic expression profiling across the pediatric systemic inflammatory response syndrome, sepsis, and septic shock spectrum," *Crit Care Med*, vol. 37, no. 5, pp. 1558–1566, May 2009, doi: 10.1097/CCM.0b013e31819fcc08. Dataset GSE13904.**

*Bioinformatics tools used to identify cellular pathways (references in publication)*

- D.A.V.I.D. (Database for Annotation, Visualization and Integrated Discovery
- PANTHER Classification System
- ToppGene

<u>*Results data analysis*</u>

*Functional annotation at day 1 sepsis:*

PANTHER

- Pathway: T-cell activation (5.1E–15)

- Biological process: MHC II-mediated immunity (1.8E–17)
- Molecular function: major histocompatibility complex antigen (5.5E–14)

ToppGene

- Molecular function: MHC class II receptor activity (<1.0E–6)
- Biological process: antigen processing and presentation (<1.0E–6)
- Mouse phenotype: abnormal antigen processing via MHC class II (<1.0E–6)
- Pathway: antigen processing and presentation (<1.0E–6)

D.A.V.I.D.

- MHC class II receptor activity (1.6E–9)

*Functional annotation at day 3 sepsis:*

PANTHER

- Pathway: T-cell activation
- Biological process: T-cell–mediated immunity
- Molecular function: major histocompatibility complex antigen

ToppGene

- Molecular function: MHC class II receptor activity
- Biological process: antigen processing via MHC class II
- Mouse phenotype: abnormal immune system physiology
- Pathway: antigen processing and presentation

D.A.V.I.D.

- MHC class II receptor activity

*Conclusions from publication-abstract:*

- septic shock: repression of genes corresponding to adaptive immunity and zinc-related biology.
- Causality remains to be elucidated.

**T. P. Shanley *et al.*, "Genome-level longitudinal expression of signaling pathways and gene networks in pediatric septic shock," *Mol Med*, vol. 13, no. 9–10, pp. 495–508, Oct. 2007, doi: 10.2119/2007-00065. Dataset GSE8121**

*Bioinformatics tools for pathway analysis*

- Ingenuity
- D.A.V.I.D.
- PANTHER

*Results data analysis*

Ingenuity Pathways Analysis, upregulated genes:

Day 1 and day 3 septic shock:

- B Cell Receptor Signaling
- GMCSF signaling
- IL10 signaling

- NFkB signaling
- p38 MAPK signaling
- Complement and coagulation
- TLR signaling
- IL6 signaling

Only day 3 septick shock:

- Integrin signaling
- IGF1 signaling
- Insulin Receptor Signaling
- PPAR signaling

*Ingenuity Pathways Analysis, downregulated genes*

- Antigen presentation
- NK cell signaling
- T cell receptor signaling

PANTHER Gene Networks, upregulated genes:

Only day 1 sepsis:

- Oxidative stress response
- Inflammation mediated by chemokine and cytokine signaling
- PDGF signaling

Only day 3 sepsis

- Inflammation mediated by chemokine and cytokine signaling
- Toll receptor signaling pathway
- Plasminogen activating cascade
- PDGF signaling pathway
- Insulin/IGF pathway-protein kinase B signaling pathway

PANTHER Gene Networks, downregulated genes:

Only day 1 sepsis:

- T cell signaling
- Wnt signaling pathway

Only Day 3 sepsis:

- T cell activation
- B cell activation

*Conclusions from publication- abstract:*

- Differential regulation of genes involved in multiple signaling pathways and gene networks primarily related to immunity and inflammation.
- distinct gene networks involving T cell- and MHC antigen-related biology were downregulated on both day one and day three.
- downregulation of genes corresponding to functional annotations related to zinc homeostasis.

General: The data further advance our genome-level understanding of pediatric septic shock and support novel hypotheses.

**N. Cvijanovich *et al.*, "Validating the genomic signature of pediatric septic shock," *Physiol Genomics*, vol. 34, no. 1, pp. 127–134, Jun. 2008, doi: 10.1152/physiolgenomics.00025.2008. Dataset GSE9692**

*Bioinformatics tools used for data analysis*

- Ingenuity Pathways Analysis
- PANTHER

*Results data analysis*

Ingenuity Pathways Analysis, upregulated genes

1. Toll-like receptor signaling
2. Interleukin-10 signaling
3. NF-κB signaling
4. Acute phase response signaling
5. p38 MAP kinase signaling
6. Complement system
7. Hepatic cholestasis
8. LXR/RXRα activation
9. Interleukin-6 signaling
10. PPARα/RXRα activation

Ingenuity Pathways Analysis, Down-regulated genes

- Natural killer cell signaling
- T-cell receptor signaling
- Antigen presentation pathway
- Interleukin-4 signaling

Gene selection from the original reported gene list [7] led to a 50 gene predictor gene classifier.

Using Ingenuity, this classifier was found to be related to:

- NF-κB signaling
- Leukocyte extravasation signaling
- Liver X receptor/Retinoid X receptor signaling
- T cell receptor signaling
- p38 MAP kinase signaling

*Conclusions from publication-abstract:*

- validation cohort: repression of genes related to zinc homeostasis and lymphocyte function

**M.-A. Cazalis *et al.*, "Early and dynamic changes in gene expression in septic shock patients: a genome-wide approach," *Intensive Care Medicine Experimental*, vol. 2, no. 1, p. 20, Aug. 2014, doi: 10.1186/s40635-014-0020-3. Dataset GSE57065**

*Bioinformatics tools used for data analysis*

- Ingenuity Pathways Analysis

*Results data analysis*

Downregulated pathways

1. Calcium-induced T lymphocyte Apoptosis
2. iCOS-iCOSL signaling in T helper cells
3. Cytotoxic T lymphocyte mediated apoptosis of target cells
4. PKCO Signalingin T lymphocytes
5. Role of NFAT in regulation of the immune response
6. Nur77 signaling in T lymphocytes
7. CD28 signaling in T helper cells
8. OX signaling pathway
9. Allograft rejection signaling
10. B cell development

Upregulated pathways:

1. Hepatic fibrosis/Hepatic stellate activation
2. P38MAPK signaling
3. IL10 signaling
4. LXR/RXR activation
5. Complement system
6. O Glycan Biosynthesis
7. NOS signaling

8. Glutathione metabolism
9. Hypoxia signaling in the cardiovascular system
10. IL-6 signaling

*Conclusions from abstract*

- both pro- and anti-inflammatory processes are induced within the very first hours after septic shock.
- more severe patients did not exhibit the strongest modulation.
- “This reinforces the idea that an immediate and tailored aggressive care of patients, aimed at restoring an appropriately regulated immune response, may have a beneficial impact on the outcome.”

**F. Venet *et al.*, “Modulation of LILRB2 protein and mRNA expressions in septic shock patients and after ex vivo lipopolysaccharide stimulation,” *Hum Immunol*, vol. 78, no. 5–6, pp. 441–450, Jun. 2017, doi: 10.1016/j.humimm.2017.03.010. Dataset GSE95233**

*Bioinformatics tools used for data analysis:* none. Differential expression analysis identified LILRB2 (leukocyte immunoglobulin-like receptors subfamily B, member 2) as higher expressed in sepsis.

*Conclusion from publication-abstract:*

- LILRB2 protein and mRNA expressions are deregulated on monocytes after septic shock

**Dataset GSE11755 (2008). No associated publication.**

Conclusion from GEO information (not published since 2008):

- Definition of an expression profile for meningococcal sepsis.
